# Red blood cell distribution width/albumin ratio is a novel risk factor of incidence and long-term mortality in chronic heart failure patients: three large cohorts from China and America

**DOI:** 10.1101/2023.09.07.23295180

**Authors:** Lin Zhang, Ying Zhou, Kaiyue Wang, Liming Wang, Tekleab Teka, Jiashun Zhou, Houliang Zhang, Xuebin Fu, Lele Zhang, Xuemei Zhang, Zhifei Fu, Lifeng Han, Xiumei Gao

## Abstract

**Aims:** Red blood cell distribution width/albumin ratio (RAR) is a novel parameter associated with inflammation. Previous studies have not focused on the role of RAR in the incidence and long-term prognosis of chronic heart failure (CHF). This study included three cohorts, two prospective and one retrospective study. The aim was to investigate the value of RAR in the incidence of CHF and the long-term prognosis of CHF.

**Methods:** Three cohorts were investigated, including MIMIC, NHANES, and JHDH. The included patients of MIMIC, NHANES, and JHDH were 22,672 from 2001-2012, 60,754 from 1999-2020, and 15,102 from 2021-2022, respectively. According to whether the patients have CHF-related risk factors, the patients were classified into non-CHF, pre-CHF, and CHF groups. The median follow-up time of MIMIC and NHANES was 364 days and 62 months. Logistic regression, Cox regression, restricted cubic spline (RCS), and Kaplan–Meier (KM) curves were used to analyze the value of RAR in CHF patients.

**Result:** In NHANES, the CHF prevalence in quartiles of RAR (Q1, Q2, Q3, and Q4) were 8.88%, 16.86%, 27.65%, and 46.61%, respectively. MIMIC and JHDH showed a similar trend. Among the non-CHF and CHF patients, the odds ratio (OR) was 1.45 (JHDH 95% CI 1.33-1.58) and 1.93 (NHANES 95% CI 1.41-2.65). In NHANES, the RAR OR value of Q2, Q3 and Q4 were 2.02(CI 1.19-3.43), 3.24(CI 1.95-5.39), and 4.95(CI 2.44-10.02) compared with Q1, respectively. And the OR was 1.05 (MIMIC 95% CI 1.02-1.07) in pre-CHF and CHF patients. The CHF mortality showed an adjusted hazard ratio (HR) is 1.12 (MIMIC 95% CI 1.1-1.14) and 2.26 (NHANES 95% CI 1.52-3.36). KM demonstrates that higher RAR (>3.4 in NHANES and >5.06 in MIMIC) prognoses lead to poor survival in CHF patients. CHF mortality in the 19th quartile of the RAR was 1.4 times higher than in the first quartile, compared with 1.22 times in the red blood cell distribution width (RDW). The 19-quartile mortality curves of the RAR were more stable than RDW and albumin (ALB).

**Conclusion:** RAR is an independent risk factor for incidence and all-cause long-term mortality in CHF patients. The predictive value of RAR for all-cause mortality in CHF is superior to ALB and RDW. RAR may be a potential clinical indicator for future treatment of CHF.

## 1. Introduction

Chronic heart failure (CHF) is the fastest-increasing cardiovascular disease, which carries poor outcomes, dramatic mortality, and high rehospitalization rates worldwide[1]. Generally, the evolution of CHF could be divided into three periods, from normal population to pre-CHF individuals and CHF patients finally[2]. Many factors (e.g., age[3] and atrial fibrillation[4]) are involved in the evolution of this process, and controlling these factors[5] can decrease or even block the development of CHF. Although many indicators had good prognoses in CHF patients, they are difficult to promote in non-CHF patients (e.g., SOFA score[6, 7], or RNA test[8]), because the relevant tests are not generally performed on non-CHF populations. Therefore, searching for clinically common and readily available predictors is essential in preventing the evolution of CHF.

Among the commonly used clinical laboratory tests, red blood cell distribution width (RDW)[9, 10] and serum albumin (ALB)[11, 12] are independent predictive factors for patients with CHF but with low odds ratio. The RDW is a risk factor for CHF, while ALB is a protective factor. Will combining the two give a better predictive value than a single in CHF? The previous study indicated that RDW and ALB ratio (RAR) was an outstanding prognostic factor of various diseases, including acute kidney injury[13], heart attack[14], and burn surgery[15]. Ni *et, al*.[16] verified that higher RAR could increase short-term mortality in heart failure (90 days).

However, there are currently three unresolved issues for RAR in CHF. Firstly, the evolution of CHF takes time, and no studies have focused on the value and relationship of RAR in CHF, pre-CHF, and CHF. Secondly, no studies have focused on the long-term predictive value of RAR in CHF patients either. Finally, the optimal cut-off value of RAR in predicting the long-term prognosis of CHF patients has also received no attention.

To sum up, RAR might play a crucial role in the progress of CHF, and relevant cohort investigation was designed to validate in the present study. This study had three objectives, i) analyze the relationship between RAR in non-CHF, pre-CHF, and CHF patients; ii) explore the optimal value of RAR to define the prognosis of CHF; iii) analyze the long-term prognosis of CHF patients at various levels of RAR. To well validate the conclusion, three separate cohorts were established, including the medical information mart for intensive care (MIMIC), the national health and nutrition examination survey (NHANES), and Jinghai district hospital (JHDH). The NHANES and JHDH were processed for non-CHF and CHF. The MIMIC was applied for the relation of pre-CHF and CHF. Furthermore, the optimal cut-off RAR value for mortality was finished in MIMIC and NHANES.

## 2. Methods

### 2.1 Study participants

Three cohorts were absorbed in this study. The NHANES and MIMIC were prospective cohort studies, while JHDH was cross-sectional. The following analysis is shown in **Figure 1**.

**Figure 1.**
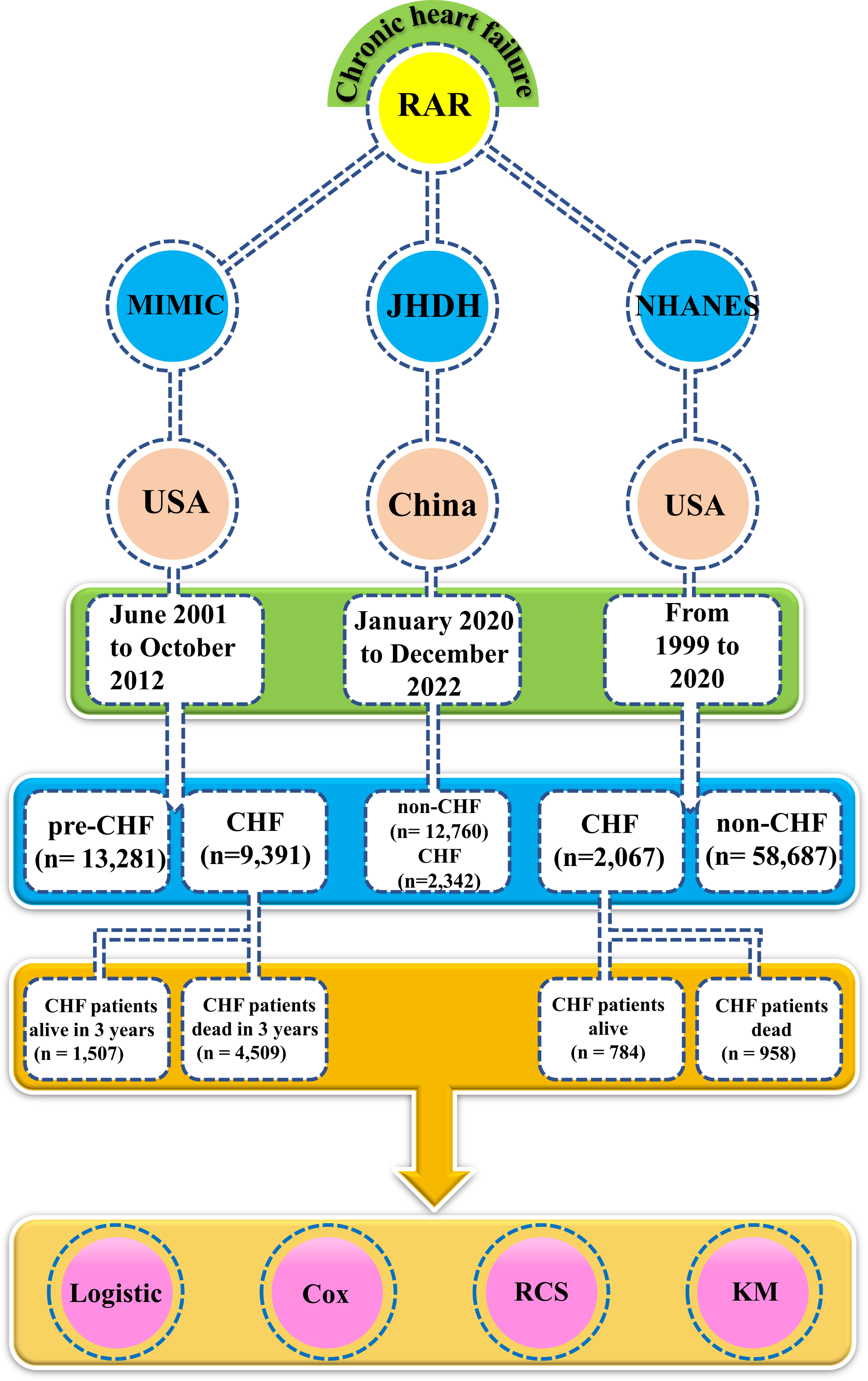
The flow chart of this study

NHANES, a periodic health-related survey in America, was processed by the Centers for disease control and Prevention’s National Center for Health Statistics. This dataset currently contained 111,797 patients and was open to the public free of charge. The dataset contained numerous survey examination data, including measurements, precisely designed health questionnaires and laboratory tests. The participants from JAN 01, 1999, to DEC 31, 2020 were included in this work.

MIMIC database (MIMIC III v1.4), a publicly single-centre critical care database, was developed by the Institutional Review Boards of Beth Israel Deaconess Medical Center and the Massachusetts Institute of Technology. The database contains charted events, laboratory events, and documents from the International Classification of Diseases (ICD-9) codes. The dataset is also free of charge, similarily to NHANES. However, users must pass a test to be eligible to register for the database and be approved by the MIMIC-III database manager. After passing the training course, the first author (Dr. Lin Zhang) was supported for extracting data from this database for the study (certification number record ID: 55394138).

In JHDH, the patients diagnosed with non-CHF and CHF in the second cardiac ward of Jinghai Hospital, according to the 2022 AHA/ACC/HFSA HF Guideline[17, 18]. This cohort aims to compare the difference in the distribution of RAR between patients in CHF and non-CHF. The hospital’s ethics committee passed the approval (JHYYLL-2022-0307).

### 2.2 The definition of heart failure

In NHANES, the diagnosis of CHF relied on a Medical Conditions Questionnaire. Only individuals who answered the questionnaire were included in the study. According to “yes” and “no”, the crowd was divided into the non-CHF or CHF group. With the CHF guideline[17, 18], CHF-related risk factors were determined, including hypertension, atherosclerotic cardiovascular disease (ASCVD), angina, and heart attack.

In MIMIC, the diagnosis of CHF relied on the ICD9 code. According to the CHF guidelines[17, 18], if patients had risk factors (e.g., hypertension, stroke, and diabetes) for CHF while they did not develop as CHF, we classified them into the pre-CHF group. We organized them into the CHF group if they had developed CHF. The diagnosis material of 131 related ICD9 codes was shown in **Table S1.**

In JHDH, the diagnosis of CHF relied on CHF guidelines[17, 18], especially on clinical symptoms, B-type natriuretic peptide (BNP), and left ventricular ejection fraction (LVEF). With clinical symptoms or signs (e.g., breathlessness and fatigue), LVEF < 50%, and BNP > 100 pg/mL, the hospitalized patients were included in the CHF or non-CHF groups.

### 2.3 CHF mortality

For NHANES, National Death Index was linked with NHANES and directly provided the mortality-related data. As previously reported[19], CHF all-cause mortality information was derived and matched with unique individuals’ IDs. Followup time was defined as the period of blood draws time to death or end of follow-up (i.e., 31 December 2019).

For MIMIC, the mortality data were directly derived from the CSV file called PATIENTS.csv, which maintained the birth information, and date of death derived from the local social security office. Surviving volunteers had not recorded specific follow-up times, and patients who died with follow-up time were included. According to whether the CHF patients stayed for more than three years, the dead CHF patients were divided into busy within 3 years or more than three years.

### 2.4 Statistical analyses

All analysis processes were finished with R software (version 4.2.2). In cleaning clinical data, the R packages *nhanesR* (version 0.9.4.1) were served for NHANES, and pgAdmin PostgreSQL (version 14.1)[20–22] was applied in MIMIC.

In the analysis of variance, if the continuous variables were distributed with Gaussian, the Student’s t-test was applied, and else Mann–Whitney U test. A chi-square test was performed to factor data (e.g., sex and ethnicity). The continuous variables were presented as means ± SDs and proportions for factor variables.

Logistic analysis was developed to explore the odds ratio (OR) in the no-CHF and CHF groups. Cox analysis was used to calculate hazard ratios (HRs) and 95% confidence intervals (CIs) of RAR for CHF all-cause mortality. Time-to-death among groups is presented with Kaplan–Meier curves. Importantly, to optimize the cut-off for RAR in CHF patient mortality, restricted cubic spline (RCS)[23] was applied in this work. The RCS model (with 3-8 knots) examined the optimal RAR value association with CHF mortality. Only a two-tailed *P*-value of <0.05 was considered statistically significant.

## 3. Result

### 3.1 General characteristics

60,754 participants were included in the NHANES cohort, including 29,220 (48.1%) males, 31,534 (51.9%) females, with an average age of 49.8, and 19,480 (32.1%) were over 60 years old. The NHANES cohort included non-CHF (*n*=58,687) and CHF (*n*=2,067) individuals, which could represent 218,996,994 Americans (**Table 1**) according to the weights. The RAR level in CHF groups (3.49±0.02) is higher than the non-CHF group (3.10±0.00). Among the 42 characteristics, only four indicators had no significance, including basophils percentage (BaP), alanine transaminase (ALT), aspartate transaminase (AST), and sodium (Na). The results are shown in **Table 1**.

**Table 1.**
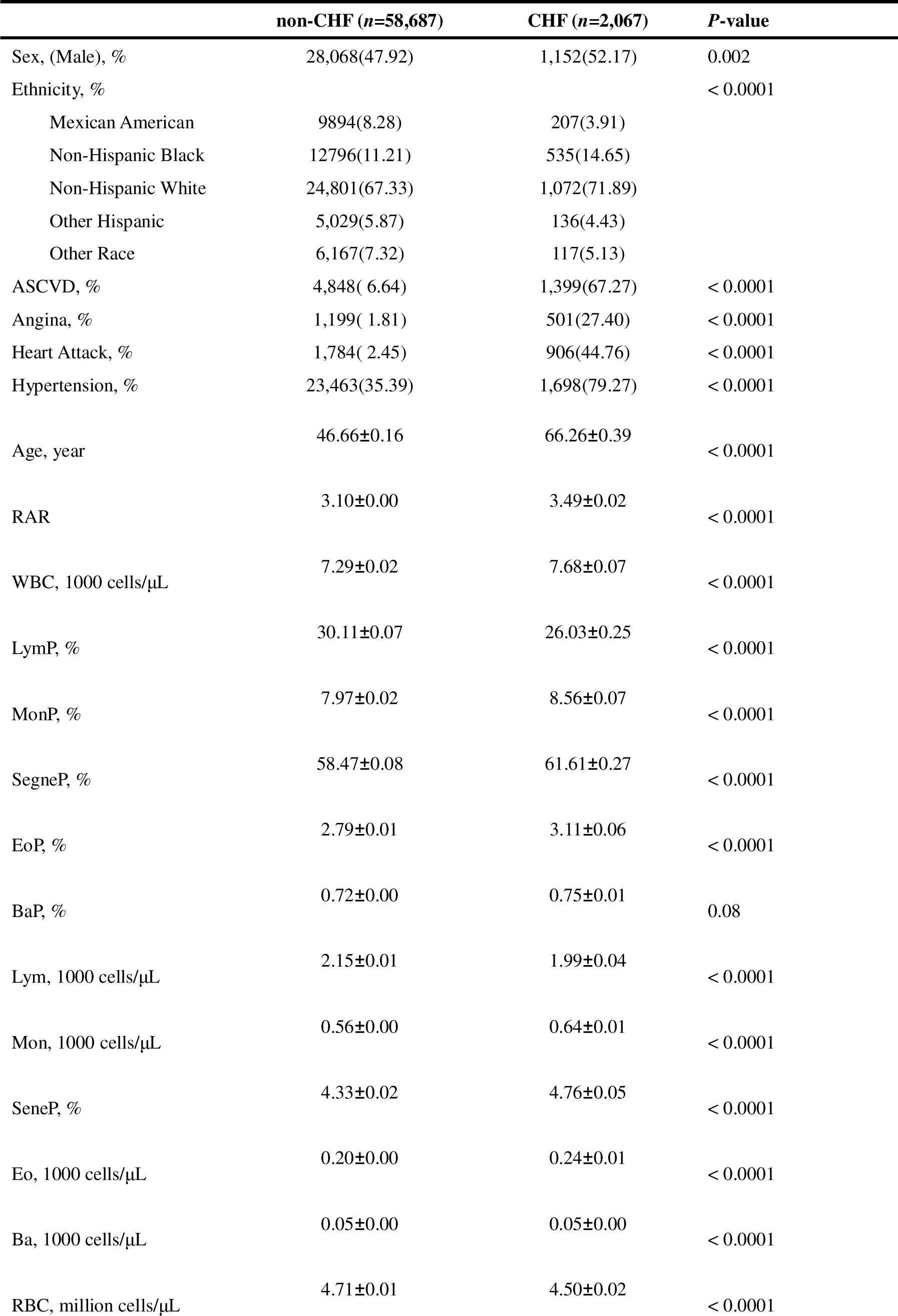
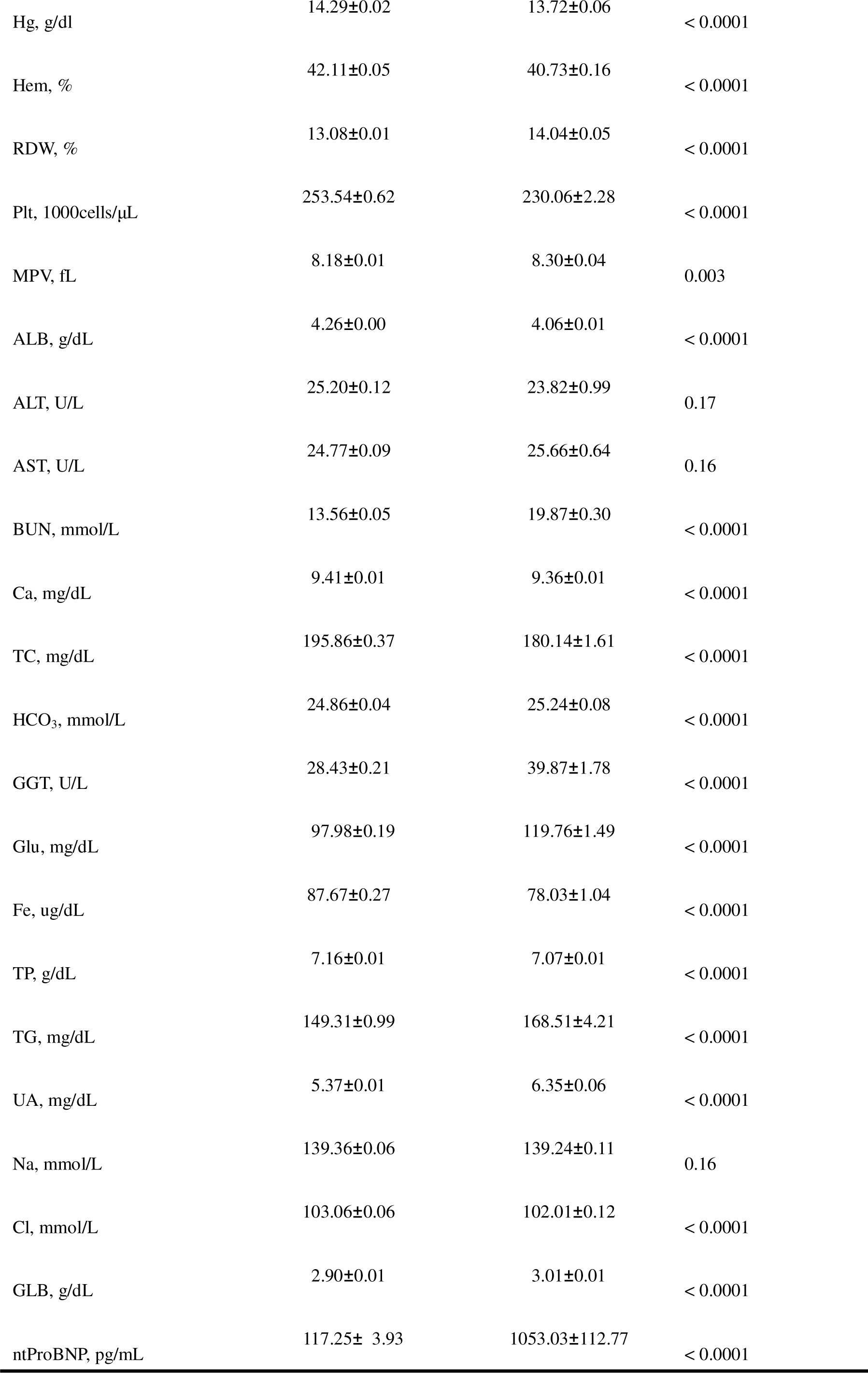
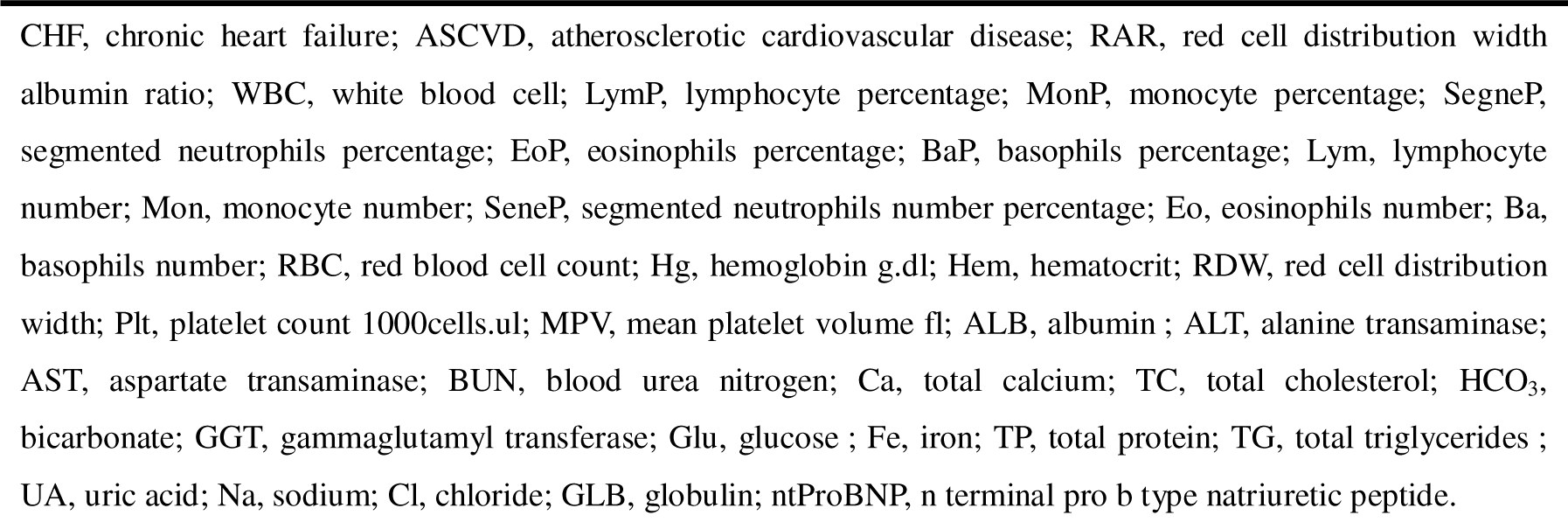
General characteristics for non-CHF and CHF in the NHANES.

22,672 participants were included in the MIMIC cohort, including 12,883 (56.8%) males, and 9,789 (43.2%) females, with an average age of 68.3, and 16,123 (71.1%) were over 60 years old. The MIMIC cohort included pre-CHF (*n*=13,281) and CHF (*n*=9,391) individuals (**Table 2**). The RAR level in CHF groups (5.08 ± 1.52) is higher than the pre-CHF group (4.74 ± 1.59). According to the results shown in **Table 2**, only four characteristics had shown no significant difference, including diabetes, platelet count (Pla), white blood cells (WBC), and anion gap (AG).

**Table 2.** General characteristics for pre-CHF and CHF in the MIMIC.

|  | **pre-CHF(n=13,281)** | **CHF (n=9,391)** | **P-value** |
| --- | --- | --- | --- |
| Age, year | 65.66 ± 14.79 | 71.86 ± 13.49 | < 0.0001 |
| Sex, (Male), % | 7841(59.04) | 5042(53.69) | < 0.0001 |
| Hypertension, % | 2391(18) | 3560(37.91) | < 0.0001 |
| Diabetes, % | 5278(39.74) | 3719(39.6) | 0.841 |
| CRD, % | 5449(41.03) | 5362(57.1) | < 0.0001 |
| CKD, % | 4747(35.74) | 5221(55.6) | < 0.0001 |
| CAD, % | 7551(56.86) | 7895(84.07) | < 0.0001 |
| RDW, % | 14.9 ± 2.08 | 15.68 ± 2.25 | < 0.0001 |
| ALB, g/dL | 3.34 ± 0.71 | 3.24 ± 0.63 | < 0.0001 |
| RAR | 4.74 ± 1.59 | 5.08 ± 1.52 | < 0.0001 |
| AG, mEq/L | 16.02 ± 5.16 | 16.07 ± 4.29 | 0.381 |
| HCO<sub>3</sub>, mEq/L | 23.81 ± 5.13 | 24.87 ± 5.42 | < 0.0001 |
| Cl, mEq/L | 102.44 ± 6.69 | 101.41 ± 6.61 | < 0.0001 |
| SCR, mg/dL | 1.68 ± 1.95 | 1.98 ± 1.9 | < 0.0001 |
| Glu, mg/dL | 165.62 ± 122.08 | 155.76 ± 90.52 | < 0.0001 |
| K, mEq/L | 4.32 ± 0.91 | 4.47 ± 0.94 | < 0.0001 |
| Na, mEq/L | 137.9 ± 5.4 | 137.82 ± 5.33 | 0.321 |
| BUN, mg/dL | 28.28 ± 22.61 | 37.61 ± 26.36 | < 0.0001 |
| HCT, % | 35.15 ± 6.57 | 34.01 ± 6.26 | < 0.0001 |
| Hg, g/dL | 11.85 ± 2.31 | 11.26 ± 2.17 | < 0.0001 |
| INR | 1.48 ± 1.63 | 1.84 ± 2.29 | < 0.0001 |
| MCH, pg | 30.37 ± 2.67 | 29.92 ± 2.82 | < 0.0001 |
| MCHC, % | 33.72 ± 1.63 | 33.09 ± 1.65 | < 0.0001 |
| MCV, fL | 90.16 ± 7.13 | 90.46 ± 7.37 | 0.003 |
| Pla, 1000 cells/μL | 246.95 ± 126.85 | 247.86 ± 126.61 | 0.591 |
| PT, seconds | 15.47 ± 8.39 | 17.77 ± 11.49 | < 0.0001 |
| PTT, seconds | 34.83 ± 21.52 | 37.55 ± 23.7 | < 0.0001 |
| RBC, m/uL | 3.92 ± 0.78 | 3.78 ± 0.74 | < 0.0001 |
| WBC, 1000cells/μL | 11.59 ± 11.77 | 11.76 ± 9.47 | 0.221 |
| Ca, mg/dL | 8.56 ± 0.99 | 8.61 ± 0.92 | 0.002 |
| Mg, mg/dL | 1.94 ± 0.5 | 2.01 ± 0.46 | < 0.0001 |
| Pho, mg/dL | 3.66 ± 1.44 | 3.95 ± 1.43 | < 0.0001 |

CRD, chronic respiratory disease; CKD, chronic kidney diseases; CAD, coronary atherosclerotic cardiopathy; RDW, red cell distribution width; ALB, albumin; red cell distribution width albumin ratio; AG, anion gap; HCO<sub>3</sub>, bicarbonate; Cl, chloride; SCR, creatinine; Glu, glucose; K, potassium; Na, sodium; BUN, blood urea nitrogen; HCT, hematocrit; Hg, hemoglobin; INR, international normalized ratio; MCH, mean corpuscular hemoglobin; MCHC, mean corpuscular hemoglobin concentration; MCV, mean corpuscular volume; Pla, platelet count; PT, prothrombin time; PTT, partial thromboplastin time; RBC, red blood cells; WBC, white blood cells; Ca, calcium; Mg, magnesium; Pho, phosphate.

15,102 participants were included in the JHDH cohort, including 7,271 (48.1%) males, and 7,831 (51.9%) females, with an average age of 66.3, and 10,725 (71.0%) were over 60 years old. The JHDH cohort included non-CHF (*n*=12,760) and CHF (*n*=2,342) individuals (**Table S2**). The RAR level in CHF groups (4.07 ± 1.08) is higher than the non-CHF group (3.44 ± 0.81). Among the total 38 characteristics, only four had no significant difference, including monocyte percentage (MonP), monocyte (Mon), basophil count (Ba), and total bile acid (TBA).

Finally, the percentage of CHF in four quartiles of RAR was summarised in **Table 3**. In NHANES, the prevalence of CHF in Q1, Q2, Q3, and Q4 were 8.88%, 16.86%, 27.65%, and 46.61%. In MIMIC, these were 27.57%, 43.62%, 47.05%, and 47.49%. In JHDH, these were 4.75%, 9.04%, 17.42%, and 30.68%. All three cohorts indicated a higher RAR value means a significantly higher prevalence of CHF.

**Table 3.**
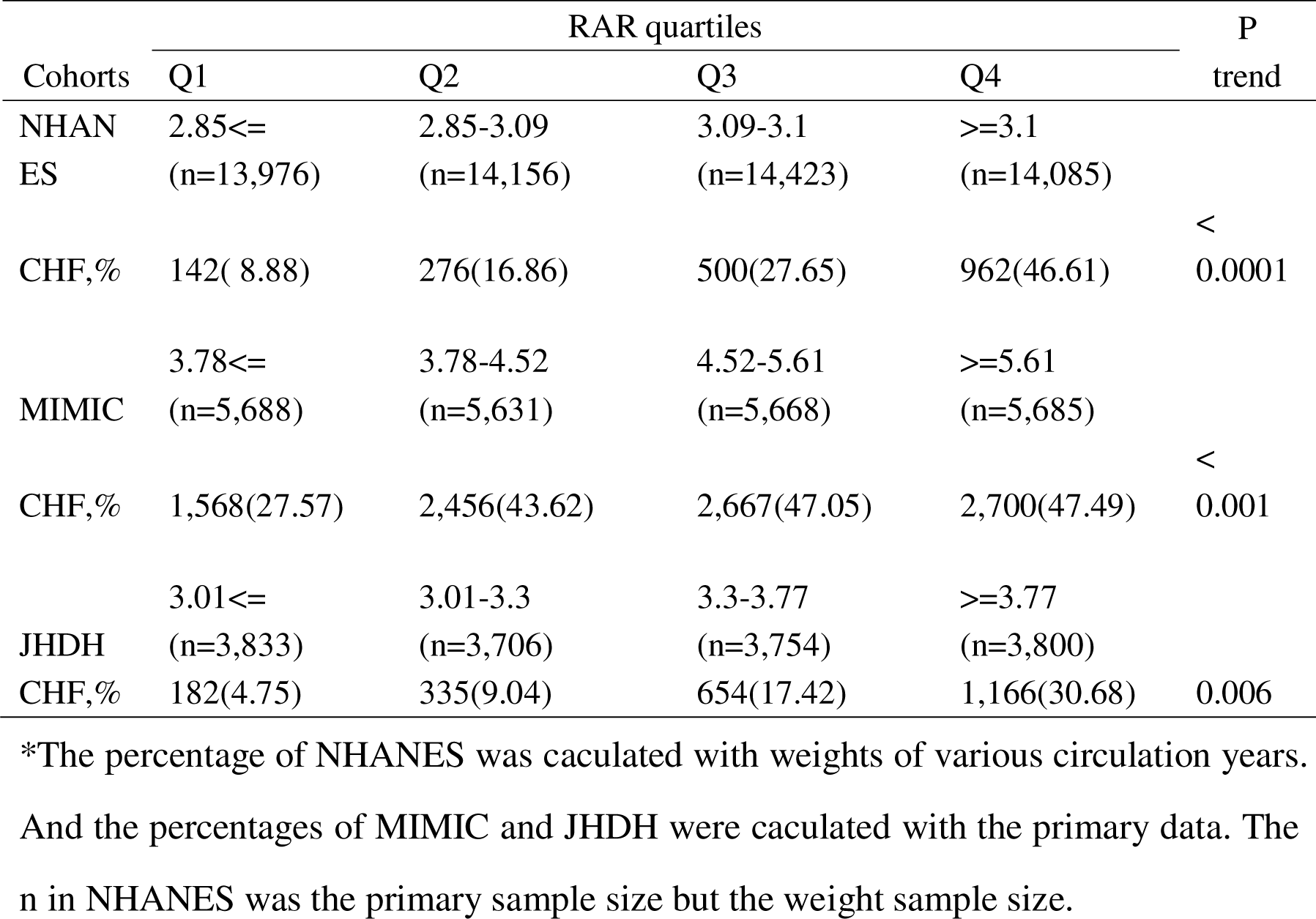
The percentage of CHF in four quartiles of RAR.

### 3.2 Logistic regression

Univariate and adjusted Logistic congression were processed among the above differential clinical features in the three cohorts (**Figure 2**). The univariate Logistic model indicated the OR in MIMIC, JHDH, and NHANES were 1.15; 95% CI 1.13-1.17, 1.87; 95% CI 1.79-1.96, and 95% CI 2.65; 2.42-2.91, respectively.

**Figure 2.**
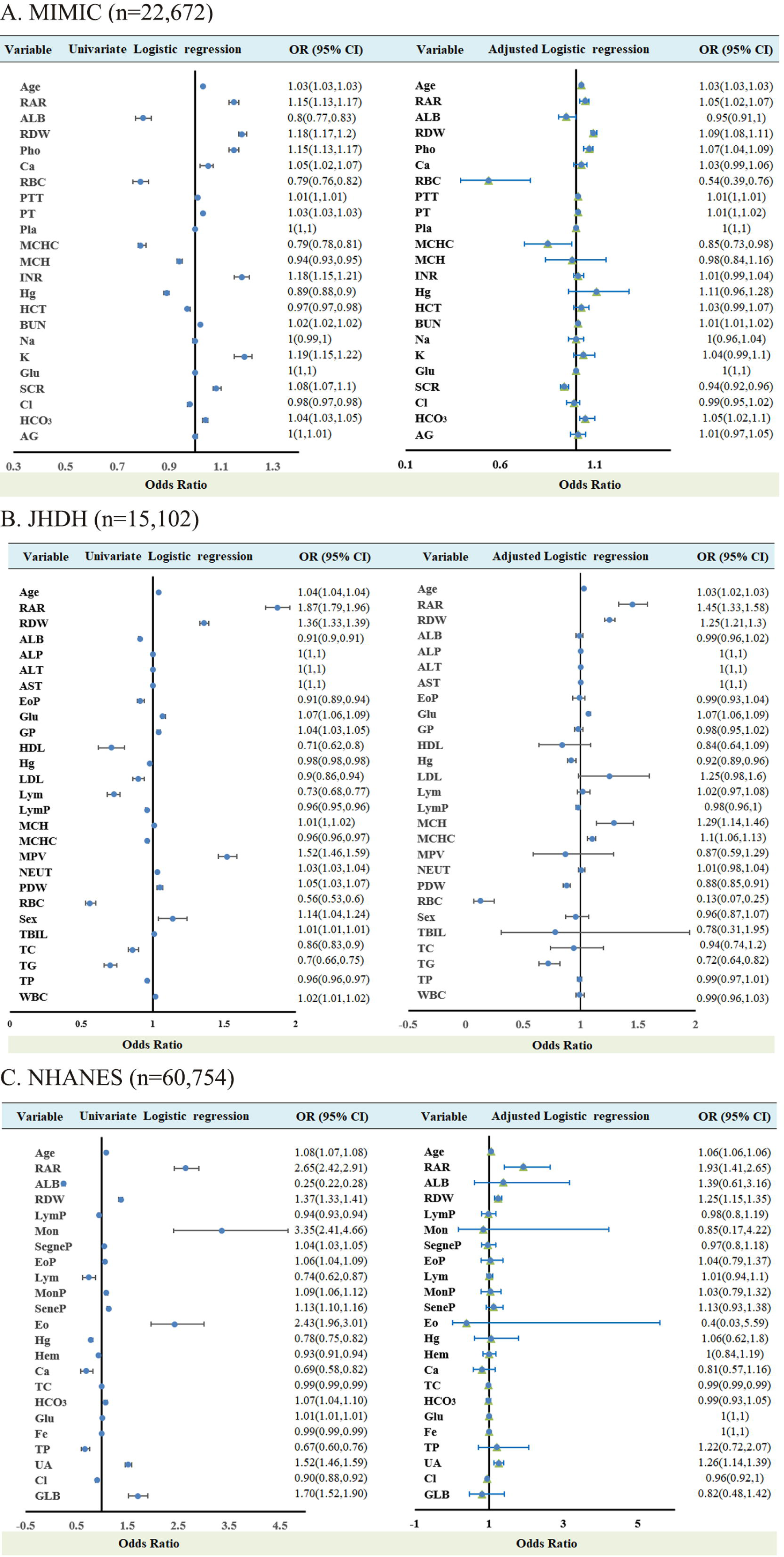
The univariate and adjusted Logistic regression. (A) The MIMIC cohort in non-CHF and CHF groups. (B) The JHDH cohort in non-CHF and CHF groups. (C) The NHANES cohort in pre-CHF and CHF groups.

The adjusted model was processed. In MIMIC, the RAR was adjusted by age, Pho, Ca, RBC, PTT, PT, Pla, MCHC, MCH, INR, Hg, HCT, BUN, Na, K, Glu, SCR, Cl, HCO_3_, and AG. Because RAR was calculated by ALB and RDW, both variables were not included to adjust RAR to eliminate overfitting. In JHDH, the RAR was adjusted by age, ALT, AST, ALP, EoP, Glu, GP, HDL, Hg, LDL, Lym, LymP, MPV, NEUT, MCH, MCHC, PDW, RBC, TC, TG, and TP. In NHANES, the RAR was adjusted by age, LymP, Mon, SegneP, EoP, Lym, MonP, SeneP, Eo, Hg, Hem, Ca, TC, HCO_3_, Glu, Fe, TP, UA, Cl, and GLB.

Among the three cohorts with adjusted, RAR was the highest risk factor associated with non-CHF and CHF patients, JHDH (OR:1.45; 95% CI 1.33-1.58) and NHANES (OR:1.93; 95% CI 1.41-2.65). And in pre-CHF and CHF groups, RAR was still a risk factor, MIMIC (OR:1.05; 95% CI 1.02-1.07). More importantly, in the included element, the adjusted RAR was the highest risk factor in two cohorts, JHDH and NHANES.

The Logistic regression for OR values at the three centres according to the quartiles of RAR are shown in **Table 4**. The Q1 was applied as a reference. In NHANES, the OR value of Q2, Q3 and Q4 were 2.02(1.19,3.43), 3.24(1.95,5.39), and 4.95(2.44,10.02), respectively. In MIMIC, the OR value of adjusted Q2, Q3, and Q4 were 1.62(1.49,1.77), 1.68(1.54,1.84), and 1.59(1.44,1.75), respectively. In JHDH, the adjusted OR value of Q2, Q3, and Q4 were 1.7(1.39,2.07), 2.99(2.44,3.65), and 5.15(4.04,6.57), respectively. The three cohorts indicated that a higher level of RAR shows a higher OR value.

**Table 4.**
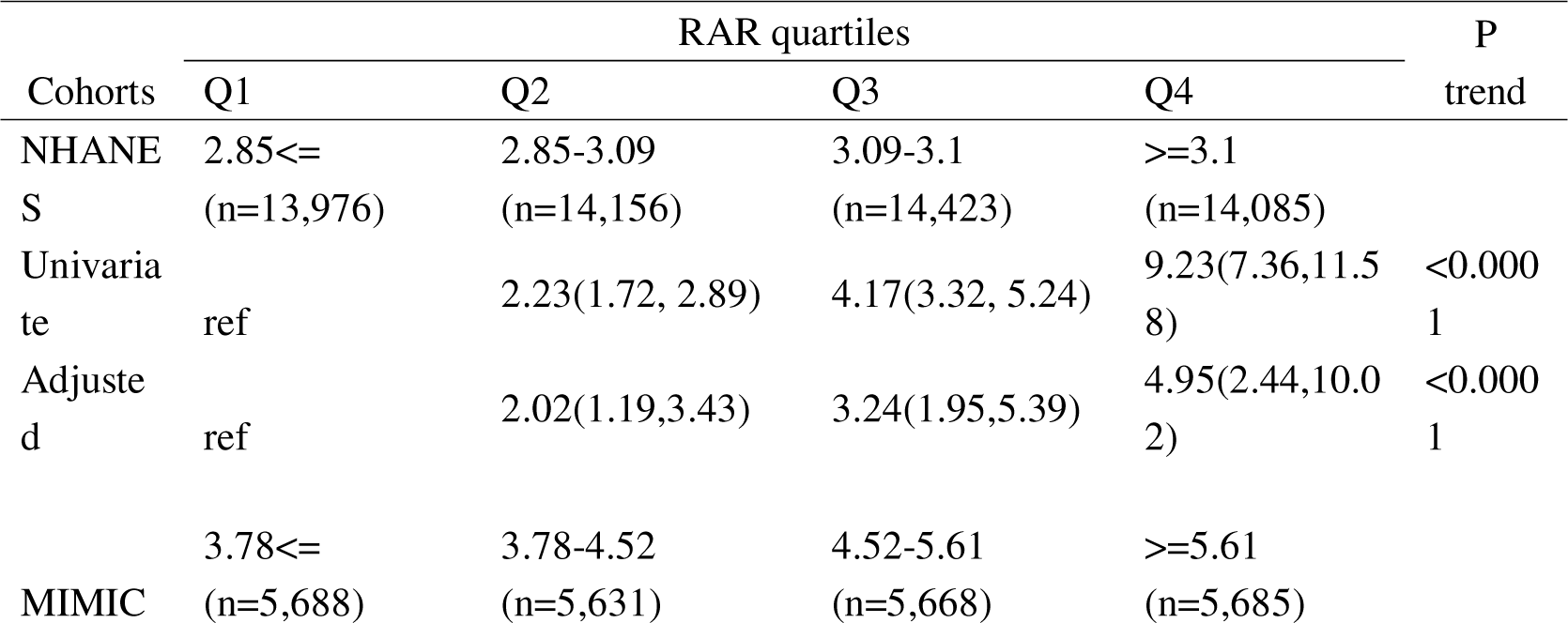
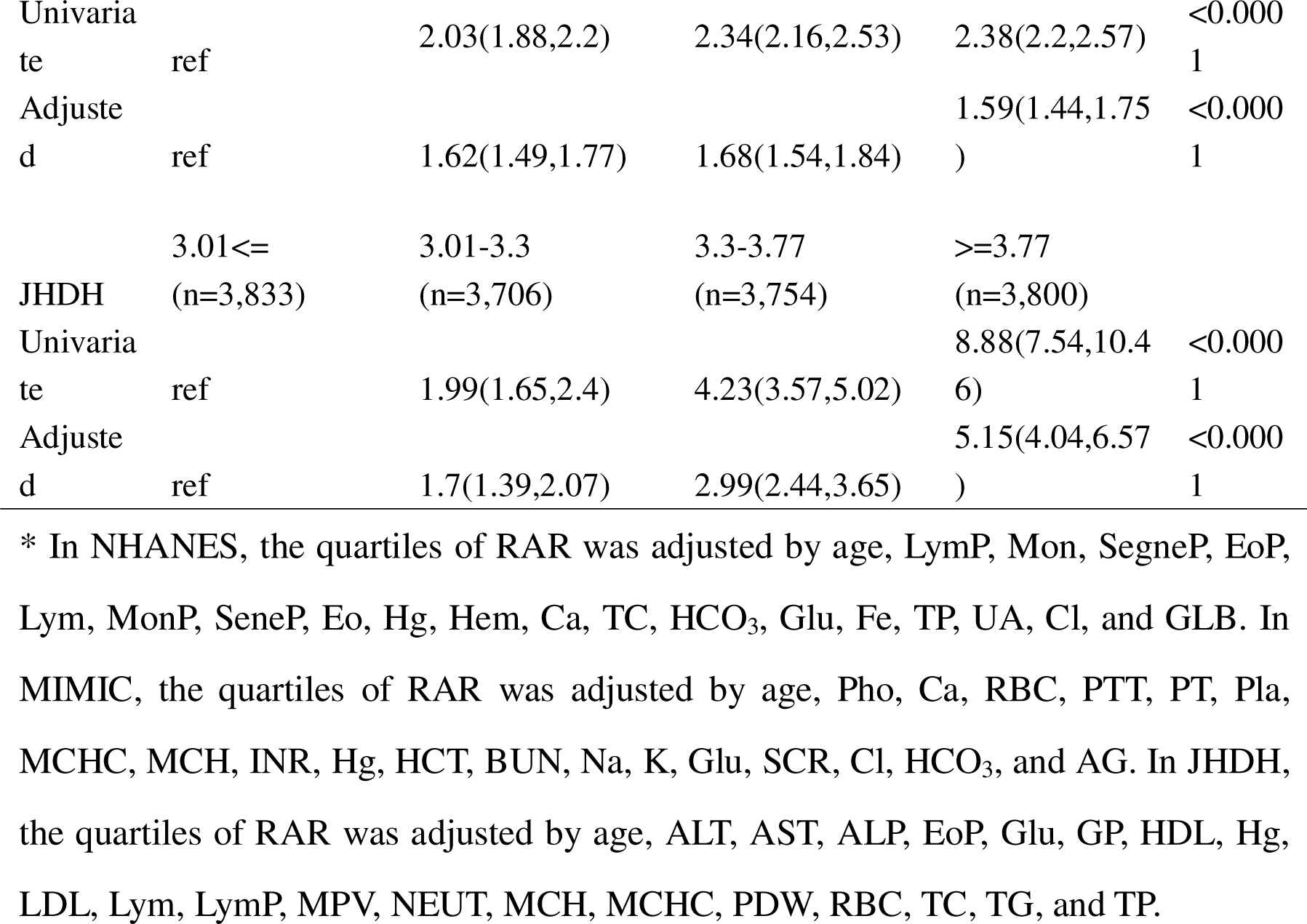
The OR value of quartiles of RAR.

### 3.3 The Cox regression

With the above significant factor of Logistic regression, the univariate and adjusted Cox regression (**Figure 3**) in MIMIC and NHANES was developed. In NHANES, all 1,742 CHF alive (*n*=784) and dead (*n*=958) in the follow-up survey (**Table S3**) had an average age of 68 years, with 965 males (55.4%). And in the MIMIC, the 6,016 CHF patients alive in 3 years (*n*=1,507) or not (*n*=4,509) had an average of 74 years (**Table S4**), with 3,208 males (53.3%). More importantly, the adjusted hazard ratio (HR) (adjusted by the same factors in the logistic analysis) in MIMIC and NHANES (**Figure 3**) was 1.12 (95% CI 1.1-1.14) and 2.26 (95% CI 1.52-3.36).

**Figure 3.**
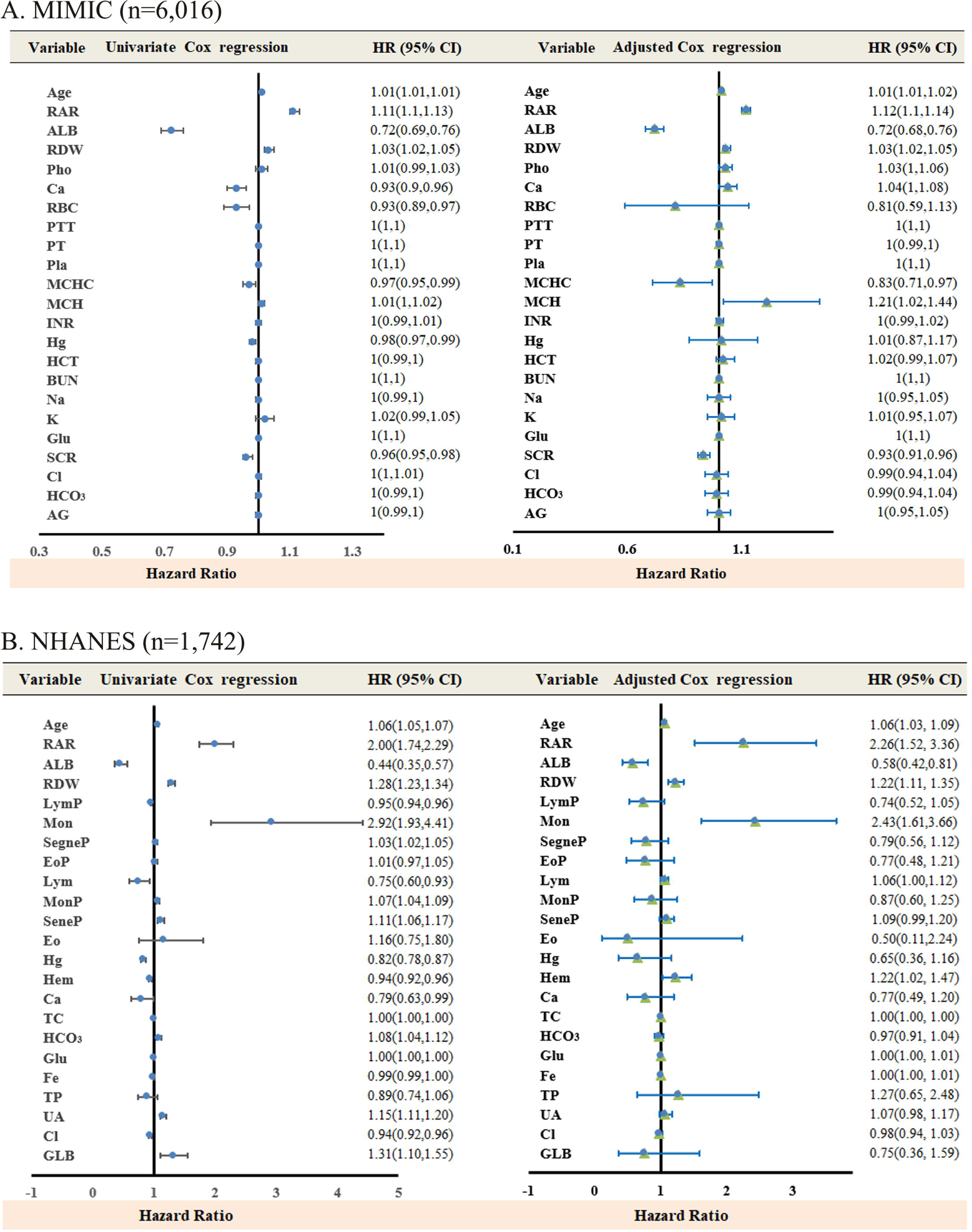
The univariate and adjusted Cox regression. (A) The CHF dead or not within 3 years in the MIMIC cohort. (B) The CHF is dead or not in the follow-up survey in the NHANES cohort.

### 3.4 RCS analysis for the optimal cut-off value

In RCS analysis, MIMIC adjusted the RAR with age, ALB, RDW, MCH, and MCHC (**Figure S1**). In NHANES, the RAR was adjusted by age, Hem, RDW, and Glu (**Figure S2**). According to the minimum fluctuation HR value range of 95% CI, the optimal value of RAR, ALB, and RDW was determined (**Figure 4**). Though the optimal cut-off value in MIMIC and NHANES differed, the trend is similar. For example, though the cut-off values of RAR are different, 5.06 in MIMIC or 3.4 in NHANES, the increasing RAR indicated a gradually rising HR. And in the ALB, both the MIMIC and NHANES showed a similarly fluctuant trend of HR.

**Figure 4.**
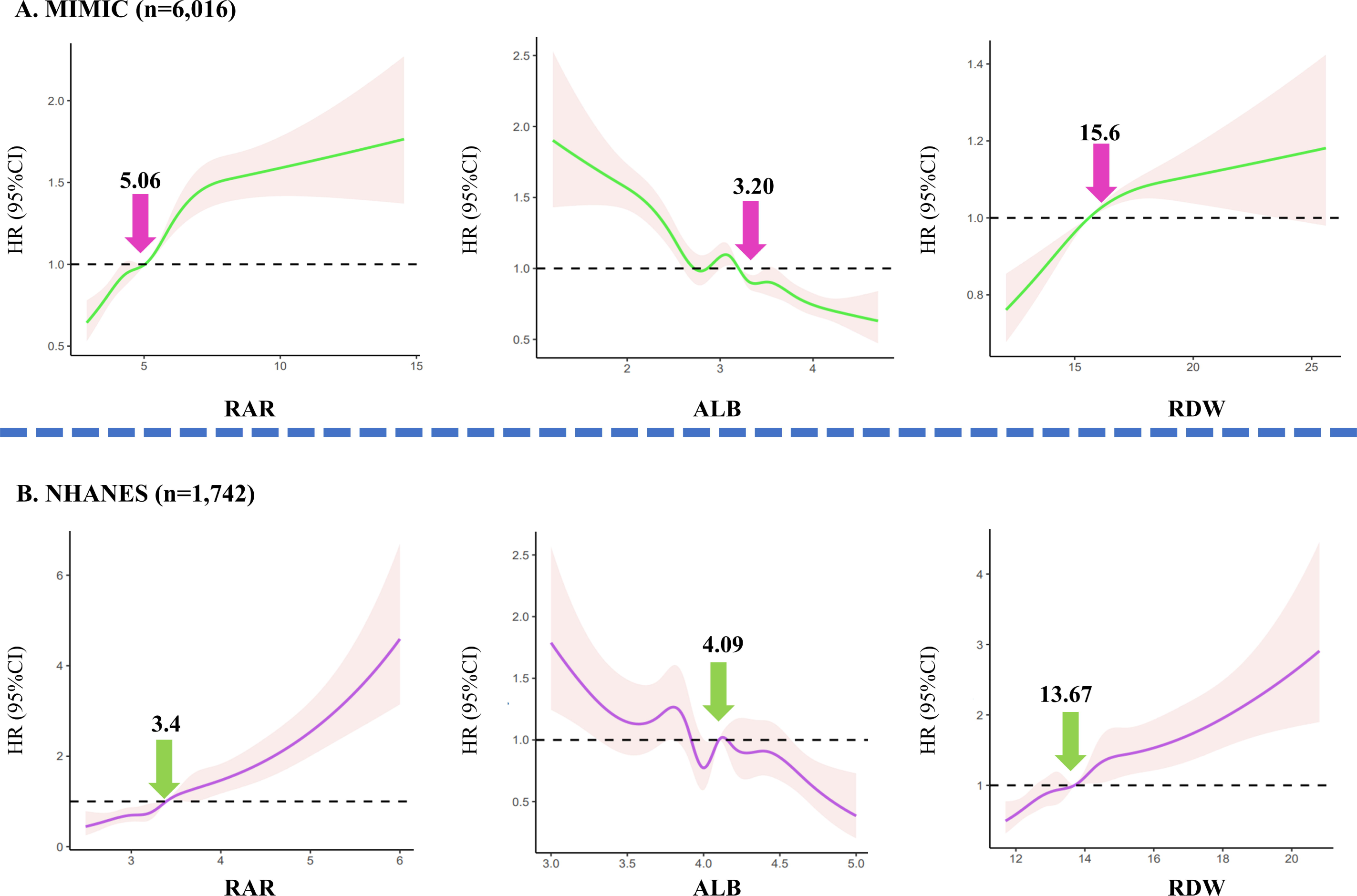
The RCS analysis for RAR, ALB, and RDW in MIMIC and NHANES cohorts.

The three factors were levels according to the optimal value of RAR, ALB, and RDW (**Figure 4**). For example, an individual in MIMIC with a RAR >5.06 was divided into the high RAR group, else the low RAR group. The RDW and ALB classified methods were similar to RAR.

### 3.5 KM survival curve

With the dichotomous RAR, ALB, and RDW levels, the KM curve was used to observe the CHF-related follow-up population. More importantly, CHF patients with RAR > 3.4 in NHANES or 5.06 in MIMIC showed a poor survival rate than the others (**Figure 5**). And the CHF patients with ALB < 4.09 in NHANES or 3.20 in MIMIC exhibited a poor survival rate than the others (**Figure 5**). RDW had a similar phenomenon to RAR (**Figure S3**). The median follow-up time of MIMIC and NHANES was 364 days and 62 months.

**Figure 5.**
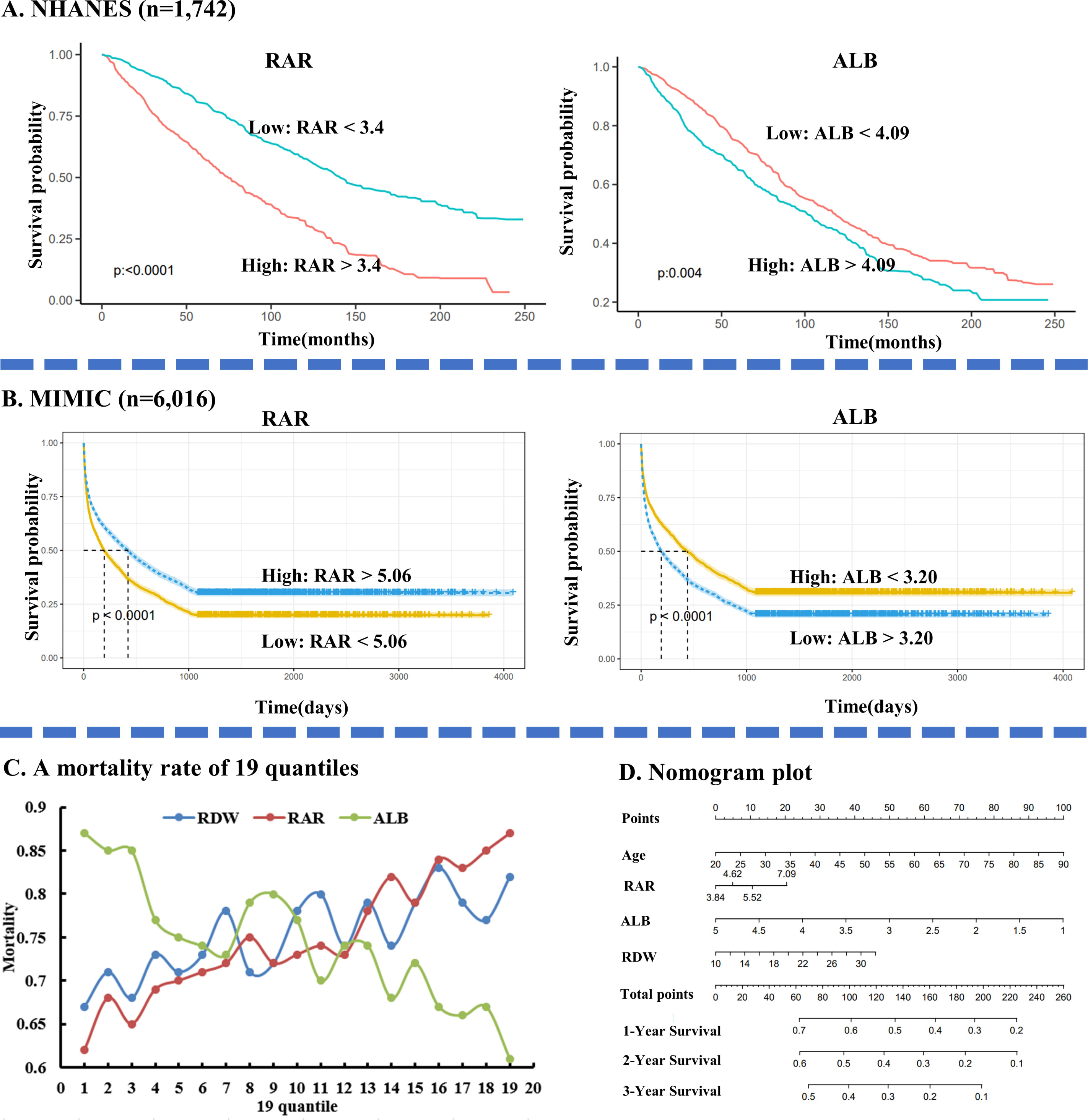
The KM curve of RAR and ALB in NHANES and MIMIC. (A) The KM curve of RAR. (B) The KM curve of ALB. (C) The mortality rate of 19 RAR, ALB, and RDW quantiles in MIMIC. (D) The nomogram plot in MIMIC.

MIMIC had more follow-up CHF patients than NHANES, which is suitable for the mortality statistics in multiple quantiles. The various quantile mortality rates in MIMIC were processed. When quantiles were set to more than 19, the 6th quantiles in ALB had a zero death population. Therefore, 19 quantiles of RAR, ALB, and RDW mortality rates were processed (**Table S5**). In the mortality of the 19 quantiles, higher RAR and RDW indicated higher mortality (**Figure 5. C**). And lower ALB demonstrated higher mortality. However, the mortality rate of RAR is more stable than RDW (**Figure 5. C**). And the mortality rate of RDW drastically fluctuated with a gradually increased. Regarding ALB, the fluctuating wave was even more drastic than RDW.

In the 1st quantile (**Table S5**), the mortality rate of RAR was lower than RDW (0.62 vs 0.67). And in the 19th quartile, the mortality rate of RAR is higher than RDW (0.87 vs 0.82). RAR’s 19th percentile mortality rate is 1.4 times higher than the 1st percentile. At the same time, it is 1.22 times for RDW. RAR was superior to RDW and ALB in predicting CHF death due to its stable predictive value at different quantile values (**Figure 5. C**). Based on the Cox analysis of CHF in MIMIC, nomogram plots were processed to predict the three-year survival of patients at RAR, RDW, ALB, and age (**Figure 5. D**).

## 4. Discussion

This multicenter study included CHF-related participants in China and America from 1999 to 2022. This study mainly answered three questions. Firstly, RAR was an independent risk factor for CHF in non-CHF or pre-CHF patients (**Figure 2**), especially for non-CHF (OR:1.93; 95% CI 1.41-2.65). Then, optimal cut-off values for RAR vary among heart failure patients (5.06 in MIMIC and 3.4 in NHANES). For example, the CHF patients in MIMIC were derived from ICU, while NHANES was community. CHF patients in MIMIC would have a much more severe condition than NHANES. Though the optimal RAR cut-off value varies, it was a good predictor of long-term CHF patient prognosis. Finally, compared with RDW and ALB, RAR had a much higher and more stable HR value in predicting the long-term prognosis of CHF patients.

RDW, a dominant hematology parameter, has been widely applied to classify various anemia[24]. Over the last decade, numerous studies confirmed that RDW is associated with ASCVSD patients, especially CHF. In a 15-year follow-up retrospective cohort with more than twenty thousand adult participants[25] without a heart attack, stroke, or CHF, a higher RDW was associated with a higher long-term incidence of CHF with an HR value of 1.47. Similarly, when RDW values were raised, a non-linear CHF incidence increased[26].

ALB was widely used to estimate the population’s nutritional status[27]. Mendelian randomization study[27] showed that lower ALB is conducive to CHF incidence with 0.54 HR. Chao *et, al*.[12] reported that ALB indicated poor short-term mortality in individuals with congestive heart failure in ICU. Similar to the RAR, ALB affects the occurrence and predicts the long-term prognosis of patients with CHF. Different from RDW, ALB is a protective factor for patients with CHF.

As an inflammation-related indicator[28], RAR received increasing attention[29–31] in recent years. However, RAR was only utilized for the short-term prognosis in diseases (30 days[30], 90 days[15, 16], 1 year[32]). For example, RAR was closely related to the incidence of contrast-related renal injury[28] and diabetic retinopathy[33]. Up to now, no studies have focused on the incidence of CHF. This is the first work to pay attention to the incidence of CHF in RAR with various cohorts. The OR (**Figure 2**) in non-CHF individuals, JHDH (1.45; 95% CI 1.33-1.58) and NHANES (1.93; 95% CI 1.41-2.65) is higher than pre-CHF individuals, MIMIC (1.05; 95% CI 1.02-1.07). In a word, RAR might better predict the progress of non-CHF to CHF patients.

In mortality, the cut-off value of RAR is different in various research. In ICU, the cut-off RAR value was 5.51[30] for cancer, 5.315 for chronic obstructive pulmonary disease[34], 4.882 for septic patients with atrial fibrillation[35], 5.5 for sepsis[31], and 5.4 for aortic aneurysms[36]. And in non-ICU wards, the cut-off value was 3.809 for patients with diabetic foot ulcers[29]. However, many previous studies have defined high and low expression of RAR by median or mean values of RAR[30]. RAR-mediated death is not necessarily linear (**Figure 5. C**), and defining high and low expression of RAR by median or mean values may not be appropriate. Therefore, we determined the optimal value of RAR by RCS. To our surprise, the optimal value of RAR to define CHF prognosis appeared to differ (5.06 in MIMIC and 3.4 in NHANES). However, the results of both cohorts were excellent, indicating that RAR is potentially valuable in clinical applications.

This study has three strengths[37], multicenter from two countries, time of duration of more than two decades and large sample size. In brief, this study consists of two prospective studies (MIMIC and NHANES) and one cross-sectional study (JHDH). The multicenter contained CHF patients derived from the community (NHANES), non-ICU wards (JHDH), and ICU (MIMIC). Furthermore, the multicenter included participants with non-CHF, pre-CHF, and CHF. However, as previously indicated[38], two limitations exist in this study. Firstly, the RAR was only detected at a single point, which may trigger the misclassification. Nevertheless, this misclassification would lead to an underestimate but not an overestimation because of the effect of regression dilution bias[39]. Secondly, the mortality in NHANES was linked with National Death Index records with a probabilistic match to result in another misclassification. Nevertheless, a previous study demonstrated that the matching method[40] had high accuracy (98.5%).

## 5. Conclusion

The higher RAR was associated with a higher incidence of CHF in non-CHF and pre-CHF individuals, especially in non-CHF. RAR could predict the long-term prognosis (over 2 decades) of CHF patients. The optimal predictive value of RAR was different for patients with varying degrees of CHF. Compared with RDW and ALB, RAR could stably predict 3-year mortality in patients with CHF.

## Data Availability

The primary MIMIC data were downloaded from https://mimic.mit.edu/. And the primary NHANES data were derived from https://www.cdc.gov/nchs/nhanes/index.htm. And the data used or analyzed in this study and the original R code are available from the corresponding author upon request.

## Acknowledgments

Thanks for the donation of the Tianjin Committee of Science and Technology of China. Thanks to Lifeng Han for proposing the idea of the article and Xiumei Gao for optimizing the frame work of the article.

## Consent for publication

This study has not been published before, and this publication has been approved by all authors.

## Ethics approval and consent to participate

The clinical trial part was approved by the Ethics Review Committee of Jinghai District Hospital.

## Authors’ contribution

LZ and YZ wrote the original draft. LZ, KYW, YL, LMW, TT, JSZ, and HLZ performed the research. LZ, YZ, XBF, and LLZ analysed the data. XMZ, ZFF, LFH, and XMG designed the experiment and revised the manuscript.

