## Supplementary material for "Red blood cell distribution width/albumin ratio is a novel risk factor of incidence and long-term mortality in chronic heart failure patients: three large cohorts from China and America": Table S1, Table S2, Table S3, Table S4, Table S5, Fig S1, Fig S2.

**Table S1.** The ICD9 item for diagnosis of CHF and pr-CHF group.

| ICD9 | Description | Group |
| --- | --- | --- |
| 40200 | Malignant hypertensive heart disease without heart failure | pre-CHF |
| 40210 | Benign hypertensive heart disease without heart failure | pre-CHF |
| 40290 | Unspecified hypertensive heart disease without heart failure | pre-CHF |
| 40400 | Hypertensive heart and chronic kidney disease, malignant, without heart failure and with chronic kidney disease stage I through stage IV, or unspecified | pre-CHF |
| 40402 | Hypertensive heart and chronic kidney disease, malignant, without heart failure and with chronic kidney disease stage V or end stage renal disease | pre-CHF |
| 40410 | Hypertensive heart and chronic kidney disease, benign, without heart failure and with chronic kidney disease stage I through stage IV, or unspecified | pre-CHF |
| 40412 | Hypertensive heart and chronic kidney disease, benign, without heart failure and with chronic kidney disease stage V or end stage renal disease | pre-CHF |
| 40490 | Hypertensive heart and chronic kidney disease, unspecified, without heart failure and with chronic kidney disease stage I through stage IV, or unspecified | pre-CHF |
| 40492 | Hypertensive heart and chronic kidney disease, unspecified, without heart failure and with chronic kidney disease stage V or end stage renal disease | pre-CHF |
| 3572 | Polyneuropathy in diabetes | pre-CHF |
| 4010 | Malignant essential hypertension | pre-CHF |
| 4011 | Benign essential hypertension | pre-CHF |
| 4019 | Unspecified essential hypertension | pre-CHF |
| 4111 | Intermediate coronary syndrome | pre-CHF |
| 4142 | Chronic total occlusion of coronary artery | pre-CHF |
| 4143 | Coronary atherosclerosis due to lipid rich plaque | pre-CHF |
| 4144 | Coronary atherosclerosis due to calcified coronary lesion | pre-CHF |
| 4148 | Other specified forms of chronic ischemic heart disease | pre-CHF |
| 4149 | Chronic ischemic heart disease, unspecified | pre-CHF |
| 4160 | Primary pulmonary hypertension | pre-CHF |
| 4372 | Hypertensive encephalopathy | pre-CHF |
| 24900 | Secondary diabetes mellitus without mention of complication, not stated as uncontrolled, or unspecified | pre-CHF |
| 24901 | Secondary diabetes mellitus without mention of complication, uncontrolled | pre-CHF |
| 24910 | Secondary diabetes mellitus with ketoacidosis, not stated as uncontrolled, or unspecified | pre-CHF |
| 24911 | Secondary diabetes mellitus with ketoacidosis, uncontrolled | pre-CHF |
| 24920 | Secondary diabetes mellitus with hyperosmolarity, not stated as uncontrolled, or unspecified | pre-CHF |
| 24921 | Secondary diabetes mellitus with hyperosmolarity, uncontrolled | pre-CHF |
| 24930 | Secondary diabetes mellitus with other coma, not stated as uncontrolled, or unspecified | pre-CHF |
| 24931 | Secondary diabetes mellitus with other coma, uncontrolled | pre-CHF |
| 24940 | Secondary diabetes mellitus with renal manifestations, not stated as uncontrolled, or unspecified | pre-CHF |
| 24941 | Secondary diabetes mellitus with renal manifestations, uncontrolled | pre-CHF |
| 24950 | Secondary diabetes mellitus with ophthalmic manifestations, not stated as uncontrolled, or unspecified | pre-CHF |
| 24951 | Secondary diabetes mellitus with ophthalmic manifestations, uncontrolled | pre-CHF |
| 24960 | Secondary diabetes mellitus with neurological manifestations, not stated as uncontrolled, or unspecified | pre-CHF |
| 24961 | Secondary diabetes mellitus with neurological manifestations, uncontrolled | pre-CHF |
| 24970 | Secondary diabetes mellitus with peripheral circulatory disorders, not stated as uncontrolled, or unspecified | pre-CHF |
| 24971 | Secondary diabetes mellitus with peripheral circulatory disorders, uncontrolled | pre-CHF |
| 24980 | Secondary diabetes mellitus with other specified manifestations, not stated as uncontrolled, or unspecified | pre-CHF |
| 24981 | Secondary diabetes mellitus with other specified manifestations, uncontrolled | pre-CHF |
| 24990 | Secondary diabetes mellitus with unspecified complication, not stated as uncontrolled, or unspecified | pre-CHF |
| 24991 | Secondary diabetes mellitus with unspecified complication, uncontrolled | pre-CHF |
| 25000 | Diabetes mellitus without mention of complication, type II or unspecified type, not stated as uncontrolled | pre-CHF |
| 25001 | Diabetes mellitus without mention of complication, type I [juvenile type], not stated as uncontrolled | pre-CHF |
| 25002 | Diabetes mellitus without mention of complication, type II or unspecified type, uncontrolled | pre-CHF |
| 25003 | Diabetes mellitus without mention of complication, type I [juvenile type], uncontrolled | pre-CHF |
| 25010 | Diabetes with ketoacidosis, type II or unspecified type, not stated as uncontrolled | pre-CHF |
| 25011 | Diabetes with ketoacidosis, type I [juvenile type], not stated as uncontrolled | pre-CHF |
| 25012 | Diabetes with ketoacidosis, type II or unspecified type, uncontrolled | pre-CHF |
| 25013 | Diabetes with ketoacidosis, type I [juvenile type], uncontrolled | pre-CHF |
| 25020 | Diabetes with hyperosmolarity, type II or unspecified type, not stated as uncontrolled | pre-CHF |
| 25021 | Diabetes with hyperosmolarity, type I [juvenile type], not stated as uncontrolled | pre-CHF |
| 25022 | Diabetes with hyperosmolarity, type II or unspecified type, uncontrolled | pre-CHF |
| 25023 | Diabetes with hyperosmolarity, type I [juvenile type], uncontrolled | pre-CHF |
| 25030 | Diabetes with other coma, type II or unspecified type, not stated as uncontrolled | pre-CHF |
| 25031 | Diabetes with other coma, type I [juvenile type], not stated as uncontrolled | pre-CHF |
| 25032 | Diabetes with other coma, type II or unspecified type, uncontrolled | pre-CHF |
| 25033 | Diabetes with other coma, type I [juvenile type], uncontrolled | pre-CHF |
| 25040 | Diabetes with renal manifestations, type II or unspecified type, not stated as uncontrolled | pre-CHF |
| 25041 | Diabetes with renal manifestations, type I [juvenile type], not stated as uncontrolled | pre-CHF |
| 25042 | Diabetes with renal manifestations, type II or unspecified type, uncontrolled | pre-CHF |
| 25043 | Diabetes with renal manifestations, type I [juvenile type], uncontrolled | pre-CHF |
| 25050 | Diabetes with ophthalmic manifestations, type II or unspecified type, not stated as uncontrolled | pre-CHF |
| 25051 | Diabetes with ophthalmic manifestations, type I [juvenile type], not stated as uncontrolled | pre-CHF |
| 25052 | Diabetes with ophthalmic manifestations, type II or unspecified type, uncontrolled | pre-CHF |
| 25053 | Diabetes with ophthalmic manifestations, type I [juvenile type], uncontrolled | pre-CHF |
| 25060 | Diabetes with neurological manifestations, type II or unspecified type, not stated as uncontrolled | pre-CHF |
| 25061 | Diabetes with neurological manifestations, type I [juvenile type], not stated as uncontrolled | pre-CHF |
| 25062 | Diabetes with neurological manifestations, type II or unspecified type, uncontrolled | pre-CHF |
| 25063 | Diabetes with neurological manifestations, type I [juvenile type], uncontrolled | pre-CHF |
| 25070 | Diabetes with peripheral circulatory disorders, type II or unspecified type, not stated as uncontrolled | pre-CHF |
| 25071 | Diabetes with peripheral circulatory disorders, type I [juvenile type], not stated as uncontrolled | pre-CHF |
| 25072 | Diabetes with peripheral circulatory disorders, type II or unspecified type, uncontrolled | pre-CHF |
| 25073 | Diabetes with peripheral circulatory disorders, type I [juvenile type], uncontrolled | pre-CHF |
| 25080 | Diabetes with other specified manifestations, type II or unspecified type, not stated as uncontrolled | pre-CHF |
| 25081 | Diabetes with other specified manifestations, type I [juvenile type], not stated as uncontrolled | pre-CHF |
| 25082 | Diabetes with other specified manifestations, type II or unspecified type, uncontrolled | pre-CHF |
| 25083 | Diabetes with other specified manifestations, type I [juvenile type], uncontrolled | pre-CHF |
| 25090 | Diabetes with unspecified complication, type II or unspecified type, not stated as uncontrolled | pre-CHF |
| 25091 | Diabetes with unspecified complication, type I [juvenile type], not stated as uncontrolled | pre-CHF |
| 25092 | Diabetes with unspecified complication, type II or unspecified type, uncontrolled | pre-CHF |
| 25093 | Diabetes with unspecified complication, type I [juvenile type], uncontrolled | pre-CHF |
| 36042 | Blind hypertensive eye | pre-CHF |
| 36211 | Hypertensive retinopathy | pre-CHF |
| 40300 | Hypertensive chronic kidney disease, malignant, with chronic kidney disease stage I through stage IV, or unspecified | pre-CHF |
| 40301 | Hypertensive chronic kidney disease, malignant, with chronic kidney disease stage V or end stage renal disease | pre-CHF |
| 40310 | Hypertensive chronic kidney disease, benign, with chronic kidney disease stage I through stage IV, or unspecified | pre-CHF |
| 40311 | Hypertensive chronic kidney disease, benign, with chronic kidney disease stage V or end stage renal disease | pre-CHF |
| 40390 | Hypertensive chronic kidney disease, unspecified, with chronic kidney disease stage I through stage IV, or unspecified | pre-CHF |
| 40391 | Hypertensive chronic kidney disease, unspecified, with chronic kidney disease stage V or end stage renal disease | pre-CHF |
| 40501 | Malignant renovascular hypertension | pre-CHF |
| 40509 | Other malignant secondary hypertension | pre-CHF |
| 40511 | Benign renovascular hypertension | pre-CHF |
| 40519 | Other benign secondary hypertension | pre-CHF |
| 40591 | Unspecified renovascular hypertension | pre-CHF |
| 40599 | Other unspecified secondary hypertension | pre-CHF |
| 41181 | Acute coronary occlusion without myocardial infarction | pre-CHF |
| 41189 | Other acute and subacute forms of ischemic heart disease, other | pre-CHF |
| 41400 | Coronary atherosclerosis of unspecified type of vessel, native or graft | pre-CHF |
| 41401 | Coronary atherosclerosis of native coronary artery | pre-CHF |
| 41402 | Coronary atherosclerosis of autologous vein bypass graft | pre-CHF |
| 41403 | Coronary atherosclerosis of nonautologous biological bypass graft | pre-CHF |
| 41404 | Coronary atherosclerosis of artery bypass graft | pre-CHF |
| 41405 | Coronary atherosclerosis of unspecified bypass graft | pre-CHF |
| 41406 | Coronary atherosclerosis of native coronary artery of transplanted heart | pre-CHF |
| 41407 | Coronary atherosclerosis of bypass graft (artery) (vein) of transplanted heart | pre-CHF |
| 41411 | Aneurysm of coronary vessels | pre-CHF |
| 40201 | Malignant hypertensive heart disease with heart failure | CHF |
| 40211 | Benign hypertensive heart disease with heart failure | CHF |
| 40291 | Unspecified hypertensive heart disease with heart failure | CHF |
| 40401 | Hypertensive heart and chronic kidney disease, malignant, with heart failure and with chronic kidney disease stage I through stage IV, or unspecified | CHF |
| 40403 | Hypertensive heart and chronic kidney disease, malignant, with heart failure and with chronic kidney disease stage V or end stage renal disease | CHF |
| 40411 | Hypertensive heart and chronic kidney disease, benign, with heart failure and with chronic kidney disease stage I through stage IV, or unspecified | CHF |
| 40413 | Hypertensive heart and chronic kidney disease, benign, with heart failure and chronic kidney disease stage V or end stage renal disease | CHF |
| 40491 | Hypertensive heart and chronic kidney disease, unspecified, with heart failure and with chronic kidney disease stage I through stage IV, or unspecified | CHF |
| 40493 | Hypertensive heart and chronic kidney disease, unspecified, with heart failure and chronic kidney disease stage V or end stage renal disease | CHF |
| 4280 | Congestive heart failure, unspecified | CHF |
| 4281 | Left heart failure | CHF |
| 4289 | Heart failure, unspecified | CHF |
| 39891 | Rheumatic heart failure (congestive) | CHF |
| 42820 | Systolic heart failure, unspecified | CHF |
| 42821 | Acute systolic heart failure | CHF |
| 42822 | Chronic systolic heart failure | CHF |
| 42823 | Acute on chronic systolic heart failure | CHF |
| 42830 | Diastolic heart failure, unspecified | CHF |
| 42831 | Acute diastolic heart failure | CHF |
| 42832 | Chronic diastolic heart failure | CHF |
| 42833 | Acute on chronic diastolic heart failure | CHF |
| 42840 | Combined systolic and diastolic heart failure, unspecified | CHF |
| 42841 | Acute combined systolic and diastolic heart failure | CHF |
| 42842 | Chronic combined systolic and diastolic heart failure | CHF |
| 42843 | Acute on chronic combined systolic and diastolic heart failure | CHF |

**Table S2.** The general characteristics of non-CHF and CHF in the JHDH

|  | non-CHF (*n*=12,760) | CHF (*n*=2,342) | *P*_value |
| --- | --- | --- | --- |
| Sex, (Male), % | 6079(47.64) | 1192(50.9) | 0.004 |
| Age, year | 65.45 ± 13.21 | 71.19 ± 12.53 | <0.0001 |
| RDW, % | 13.3 ± 1.54 | 14.45 ± 2.21 | <0.0001 |
| ALB, g/dL | 3.96 ± 0.52 | 3.67 ± 0.54 | <0.0001 |
| RAR | 3.44 ± 0.81 | 4.07 ± 1.08 | <0.0001 |
| BNP, pg/mL | 79.45 ± 72.8 | 1666.06 ± 1313.46 | <0.0001 |
| WBC, 1000 cells/μL | 8.53 ± 6.83 | 9.17 ± 6.56 | <0.0001 |
| RBC, million cells/μL | 4.25 ± 0.7 | 3.92 ± 0.89 | <0.0001 |
| Hg, g/L | 130.47 ± 22.24 | 118.22 ± 28.04 | <0.0001 |
| HCT, % | 38.81 ± 6.16 | 35.93 ± 8.13 | <0.0001 |
| MCH, fL | 91.52 ± 6.13 | 91.92 ± 7.2 | 0.011 |
| MCHC, g/L | 335.59 ± 13.6 | 328.15 ± 15.23 | <0.0001 |
| Pla, 1000 cells/μL | 232.43 ± 82.9 | 216.1 ± 103.28 | <0.0001 |
| LymP, % | 21.22 ± 10.93 | 16.71 ± 10.38 | <0.0001 |
| MonP, % | 6.89 ± 2.7 | 6.9 ± 3.24 | 0.861 |
| EoP, % | 1.37 ± 1.85 | 1.12 ± 1.89 | <0.0001 |
| Lym, 1000 cells/μL | 1.55 ± 1.53 | 1.32 ± 1.74 | <0.0001 |
| Mon, 1000 cells/μL | 0.56 ± 0.34 | 0.66 ± 3.07 | 0.141 |
| Eo, 1000 cells/μL | 0.1 ± 0.15 | 0.08 ± 0.14 | <0.0001 |
| Ba, 1000 cells/μL | 0.02 ± 0.02 | 0.02 ± 0.02 | 0.192 |
| PDW, % | 12.48 ± 2.67 | 12.84 ± 2.66 | <0.0001 |
| MPV, fL | 9.91 ± 1.03 | 10.4 ± 1.1 | <0.0001 |
| NeuP, % | 70.97 ± 12.52 | 75.7 ± 12.26 | <0.0001 |
| ALT, U/L | 26.19 ± 90.17 | 53.35 ± 231.84 | <0.0001 |
| AST, U/L | 37.27 ± 145.9 | 95.69 ± 486.34 | <0.0001 |
| ALP, U/L | 85.75 ± 48.4 | 97.62 ± 67.45 | <0.0001 |
| GGT, U/L | 41.71 ± 88.91 | 56.72 ± 82.22 | <0.0001 |
| TP, g/L | 63.99 ± 7.16 | 61.83 ± 8.07 | <0.0001 |
| GP,g/L | 24.49 ± 4.48 | 25.43 ± 4.87 | <0.0001 |
| TBIL, μmol/L | 12.47 ± 14.85 | 15.63 ± 19.13 | <0.0001 |
| DBIL, μmol/L | 5.8 ± 11.9 | 8.65 ± 15.87 | <0.0001 |
| IBIL, μmol/L | 6.67 ± 4.95 | 6.98 ± 5.9 | 0.016 |
| TBA, μmol/L | 5.2 ± 14.52 | 5.26 ± 14.3 | 0.872 |
| Glu, mg/L | 6.74 ± 2.94 | 7.54 ± 3.73 | <0.0001 |
| TG, mmol/L | 1.44 ± 1.26 | 1.17 ± 0.73 | <0.0001 |
| TC, mmol/L | 4.28 ± 1.19 | 4.07 ± 1.29 | <0.0001 |
| HDL, mmol/L | 1.16 ± 0.35 | 1.12 ± 0.36 | <0.0001 |
| LDL, mmol/L | 2.65 ± 0.98 | 2.55 ± 1.11 | <0.0001 |
| RDW, red blood cell distribution width; ALB, albumin; RAR, red cell distribution width albumin ratio; BNP, b type natriuretic peptide; WBC, white blood cells; RBC, red blood cells; Hg, hemoglobin; HCT, hematocrit; MCH, mean corpuscular volume; MCHC, mean corpuscular hemoglobin concentration; Pla, platelet; LymP, lymphocyte percentage; Monp, monocyte percentage; EoP, eosinophilic percentage; Lym, lymphocyte count; Mon, monocyte count; Eo, eosinophil count; Ba, basophil count; PDW, platelet distribution width; MPV, mean platelet volume; NeuP, neutrophil percentage; ALT, glutamic-pyruvic transaminase; AST, glutamic oxalacetic transaminase; ALP, alkaline phosphatase; GGT, glutamyltransferase; TP, total protein; GP, globulin; TBIL, total bilirubin; DBIL, direct bilirubin; IBIL, indirect bilirubin; TBA, total bile acid; Glu, glucose; TG, total triglycerides; TC, total cholesterol; HDL, high-density lipoprotein; LDL, low-density lipoprotein. | | | |

**Table S3.** The general characteristics of CHF patients who survived or died during follow-up in NHANES.

|  | CHF patients alive (*n* = 784) | CHF patients dead (*n* = 958) | *P*-value |
| --- | --- | --- | --- |
| Sex, (Male), % | 421(53.41) | 544(50.15) | 0.331 |
| ASCVD, % | 516(65.24) | 674(69.41) | 0.151 |
| Angina, % | 178(25.54) | 257(29.61) | 0.173 |
| Heart Attack, % | 332(43.47) | 439(45.99) | 0.452 |
| Hypertension, % | 662(80.11) | 769(78.21) | 0.511 |
| Ethnicity, % |  |  | < 0.001 |
| Mexican American | 89(5.11) | 103(3.14) |  |
| Non-Hispanic Black | 226(17.09) | 201(12.11) |  |
| Non-Hispanic White | 337(66.37) | 589(77.46) |  |
| Other Hispanic | 77(5.59) | 34(3.06) |  |
| Other Race | 55(5.83) | 31(4.23) |  |
| Age, year | 61.17±0.64 | 71.14±0.45 | < 0.0001 |
| RAR | 3.38±0.03 | 3.55±0.03 | < 0.0001 |
| RDW, % | 13.79±0.07 | 14.18±0.07 | < 0.001 |
| ALB, g/dL | 4.12±0.02 | 4.04±0.01 | < 0.001 |
| WBC, 1000 cells/μL | 7.59±0.11 | 7.75±0.10 | 0.291 |
| LymP, % | 28.07±0.44 | 24.25±0.40 | < 0.0001 |
| MonP, % | 8.35±0.10 | 8.70±0.11 | 0.021 |
| SegneP, % | 59.82±0.49 | 63.24±0.46 | < 0.0001 |
| EoP, % | 3.06±0.10 | 3.18±0.10 | 0.43 |
| BaP, % | 0.77±0.03 | 0.69±0.02 | 0.012 |
| Lym, 1000 cells/μL | 2.12±0.06 | 1.88±0.05 | 0.0041 |
| Mon, 1000 cells/μL | 0.61±0.01 | 0.65±0.01 | 0.011 |
| SeneP, % | 4.57±0.07 | 4.93±0.08 | 0.0021 |
| Eo, 1000 cells/μL | 0.23±0.01 | 0.24±0.01 | 0.561 |
| Ba, 1000 cells/μL | 0.06±0.00 | 0.05±0.00 | 0.003 |
| RBC, million cells/μL | 4.61±0.03 | 4.39±0.02 | < 0.0001 |
| Hg, g/dL | 13.97±0.08 | 13.49±0.08 | < 0.0001 |
| Hem, % | 41.41±0.25 | 40.06±0.23 | < 0.001 |
| Pla, 1000 cells/μL | 229.20±3.35 | 232.23±3.56 | 0.532 |
| MPV, fL | 8.42±0.07 | 8.17±0.04 | 0.0021 |
| ALT, U/L | 25.26±1.23 | 23.31±1.81 | 0.372 |
| AST, U/L | 25.94±1.05 | 26.44±0.94 | 0.721 |
| BUN, mmol/L | 16.60±0.30 | 22.86±0.53 | < 0.0001 |
| Ca, mg/dL | 9.38±0.02 | 9.36±0.02 | 0.483 |
| TC, mg/dL | 181.03±2.51 | 181.47±2.12 | 0.881 |
| HCO_3_, mmol/L | 25.07±0.11 | 25.35±0.14 | 0.112 |
| GGT, U/L | 37.30±2.47 | 40.63±2.55 | 0.341 |
| Glu, mg/dL | 117.91±2.30 | 120.58±2.15 | 0.4 |
| Fe, μg/dL | 81.38±1.82 | 75.11±1.42 | 0.011 |
| TP, g/dL | 7.05±0.02 | 7.10±0.02 | 0.191 |
| TG, mg/dL | 173.16±7.26 | 166.10±4.69 | 0.392 |
| UA, mg/dL | 6.12±0.08 | 6.65±0.09 | < 0.0001 |
| Na, mmol/L | 139.34±0.17 | 138.91±0.13 | 0.031 |
| Cl, mmol/L | 102.55±0.18 | 101.85±0.17 | 0.0041 |
| GLB, g/dL | 2.93±0.02 | 3.06±0.02 | < 0.001 |
| ntProBNP, pg/mL | 280.78±112.77 | 1340.09±152.04 | < 0.0001 |
| CHF, chronic heart failure; ASCVD, atherosclerotic cardiovascular disease; RAR, red cell distribution width albumin ratio; WBC, white blood cell; LymP, lymphocyte percentage; MonP, monocyte percentage; SegneP, segmented neutrophils percentage; EoP, eosinophils percentage; BaP, basophils percentage; Lym, lymphocyte; Mon, monocyte; SeneP, segmented neutrophils percentage; Eo, eosinophils; Ba, basophils; RBC, red blood cell; Hg, hemoglobin; Hem, hematocrit; RDW, red cell distribution width; Pla, platelet; MPV, mean platelet volume; ALB, albumin ; ALT, alanine transaminase; AST, aspartate transaminase; BUN, blood urea nitrogen; Ca, total calcium; TC, total cholesterol; HCO_3_, bicarbonate; GGT, gammaglutamyl transferase; Glu, glucose ; Fe, iron; TP, total protein; TG, triglycerides ; UA, uric acid; Na, sodium; Cl, chloride; GLB, globulin; ntProBNP, n terminal pro B type natriuretic peptide. | | | |

**Table S4.** The general characteristics of CHF patients who live more than 3 years or died within 3 years during follow-up in Mimic.

|  | CHF alive 3 years (*n*= 1,507) | CHF dead 3 years (*n*=4,509) | *P*_value |
| --- | --- | --- | --- |
| Age, year | 72.07 ± 12.24 | 75.03 ± 12.31 | < 0.0001 |
| Sex, (Male), % | 788(52.29) | 2420(53.67) | 0.368 |
| RDW, % | 15.72 ± 2.26 | 16.15 ± 2.36 | < 0.0001 |
| ALB, g/dL | 3.29 ± 0.59 | 3.09 ± 0.62 | < 0.0001 |
| RAR | 4.97 ± 1.4 | 5.49 ± 1.63 | < 0.0001 |
| Survival time, days | 1978.32 ± 656.69 | 255.35 ± 297.81 | < 0.0001 |
| Hypertension, % | 568(37.69) | 1951(43.27) | < 0.001 |
| Diabetes, % | 690(45.79) | 1641(36.39) | < 0.0001 |
| CRD, % | 815(54.08) | 2909(64.52) | < 0.0001 |
| CKD, % | 843(55.94) | 2762(61.26) | < 0.001 |
| CAD, % | 1303(86.46) | 3789(84.03) | 0.026 |
| AG, mEq/L | 16.53 ± 4.43 | 16.23 ± 4.18 | 0.024 |
| HCO_3_, mEq/L | 24.78 ± 5.4 | 24.87 ± 5.65 | 0.574 |
| Cl, mEq/L | 101.18 ± 6.58 | 101.15 ± 6.95 | 0.873 |
| SCR, mg/dL | 2.23 ± 2.14 | 2.01 ± 1.72 | < 0.001 |
| Glu, mg/dL | 161.31 ± 94.07 | 151.27 ± 83.49 | < 0.001 |
| K, mEq/L | 4.49 ± 0.92 | 4.53 ± 0.97 | 0.171 |
| Na, mEq/L | 137.94 ± 5.22 | 137.68 ± 5.77 | 0.105 |
| BUN, mg/dL | 39.99 ± 27.22 | 40.93 ± 27.32 | 0.244 |
| HCT, % | 33.91 ± 6.23 | 33.35 ± 6.06 | 0.003 |
| Hg, g/dL | 11.22 ± 2.14 | 10.97 ± 2.06 | 1.00E-04 |
| INR, | 1.99 ± 2.78 | 1.96 ± 2.6 | 0.6974 |
| MCH, pg | 29.75 ± 2.83 | 29.97 ± 2.91 | 0.0116 |
| MCHC, % | 33.08 ± 1.59 | 32.91 ± 1.63 | < 0.001 |
| MCV, fL | 89.96 ± 7.53 | 91.09 ± 7.63 | < 0.0001 |
| Pla, 1000 cells/μL | 255.33 ± 128.4 | 244.79 ± 135.35 | 0.007 |
| PT, seconds | 18.09 ± 11.41 | 18.31 ± 12.16 | 0.531 |
| PTT, seconds | 37.83 ± 24.06 | 37.71 ± 23.16 | 0.867 |
| RBC, million cells/μL | 3.79 ± 0.75 | 3.68 ± 0.72 | < 0.0001 |
| WBC, 1000 cells/μL | 11.56 ± 8.43 | 12.37 ± 11.17 | 0.003 |
| Ca, mg/dL | 8.67 ± 0.9 | 8.57 ± 0.94 | < 0.001 |
| Mg, mg/dL | 2.01 ± 0.44 | 2.02 ± 0.51 | 0.239 |
| Pho, mg/dL | 4.06 ± 1.48 | 4.05 ± 1.43 | 0.823 |
| CRD, chronic respiratory disease; CKD, chronic kidney diseases; CAD, coronary atherosclerotic cardiopathy; RDW, red cell distribution width; ALB, albumin; red cell distribution width albumin ratio; AG, anion gap; HCO_3_, bicarbonate; Cl, chloride; SCR, creatinine; Glu, glucose; K, potassium; Na, sodium; BUN, blood urea nitrogen; HCT, hematocrit; Hg, hemoglobin; INR, international normalized ratio; MCH, mean corpuscular hemoglobin; MCHC, mean corpuscular hemoglobin concentration; MCV, mean corpuscular volume; Pla, platelet; PT, prothrombin time; PTT, partial thromboplastin time; RBC, red blood cells; WBC, white blood cells; Ca, calcium; Mg, magnesium; Pho, phosphate. | | | |

**Figure S1.** The Schoenfeld individual test to adjust RAR and ALB in the NHANES cohort. (A) The beta (t) for RCS with seven knots and adjusted Age in RAR. (B) The beta (t) for RCS with eight knots and adjusted RDW in ALB.

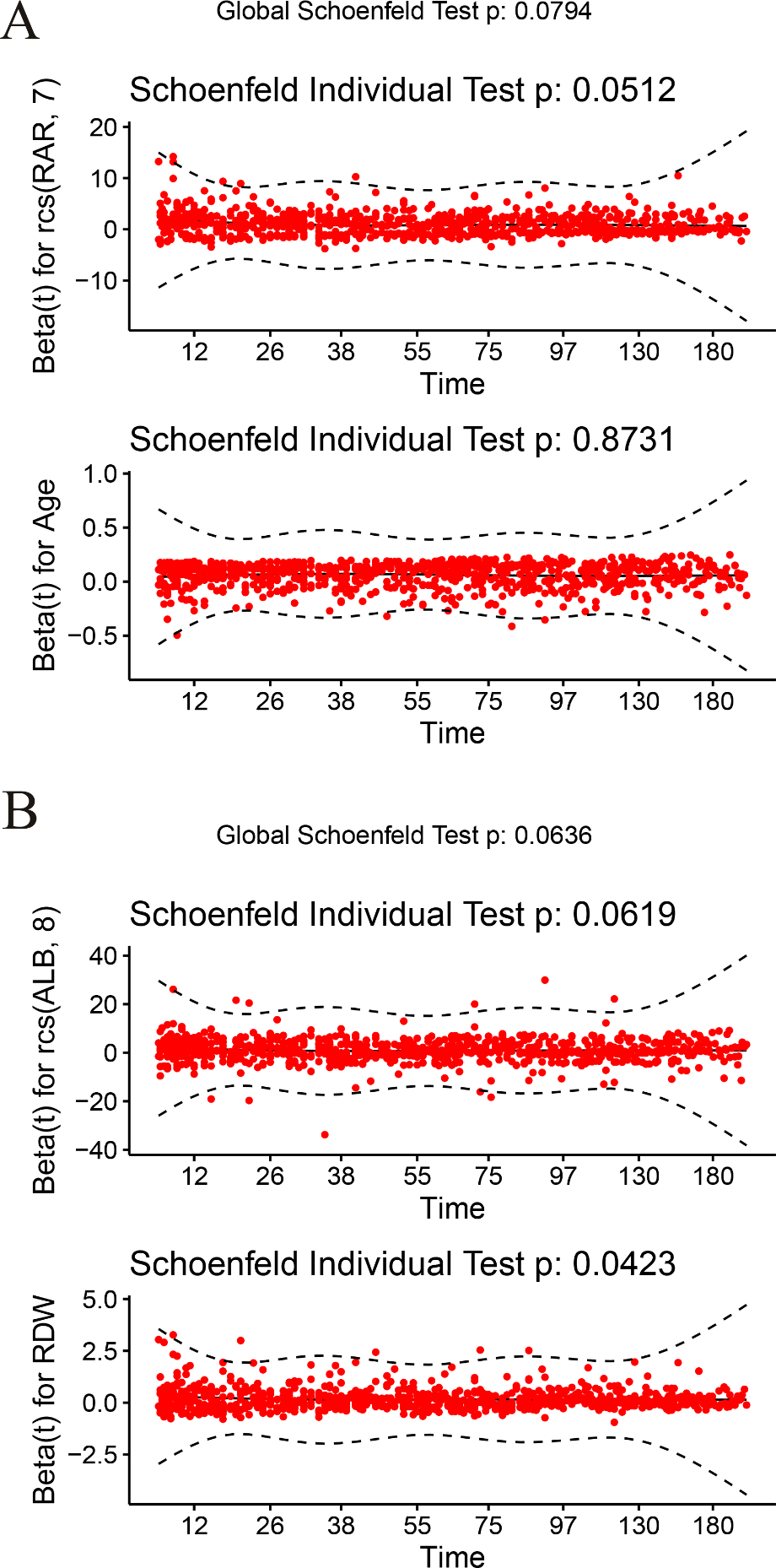

**Figure S2.** The Schoenfeld individual test to adjust RAR and ALB with MCH in the MIMIC cohort. (A) The beta (t) for RCS in RAR with five knots. (B) The beta (t) for RCS in ALB with eight knots.

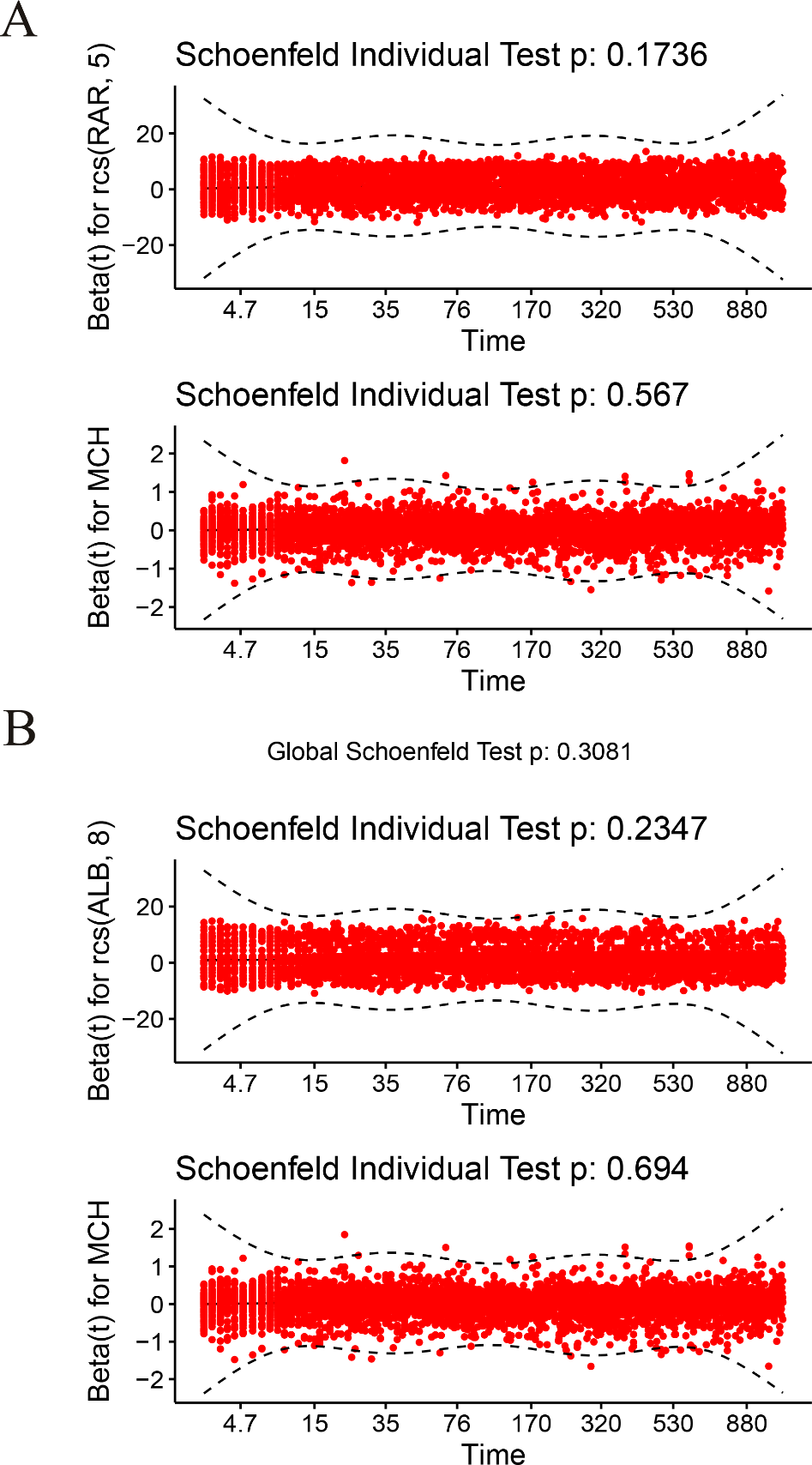

**Figure S3.** The KM curve of RDW in NHANES and MIMIC.

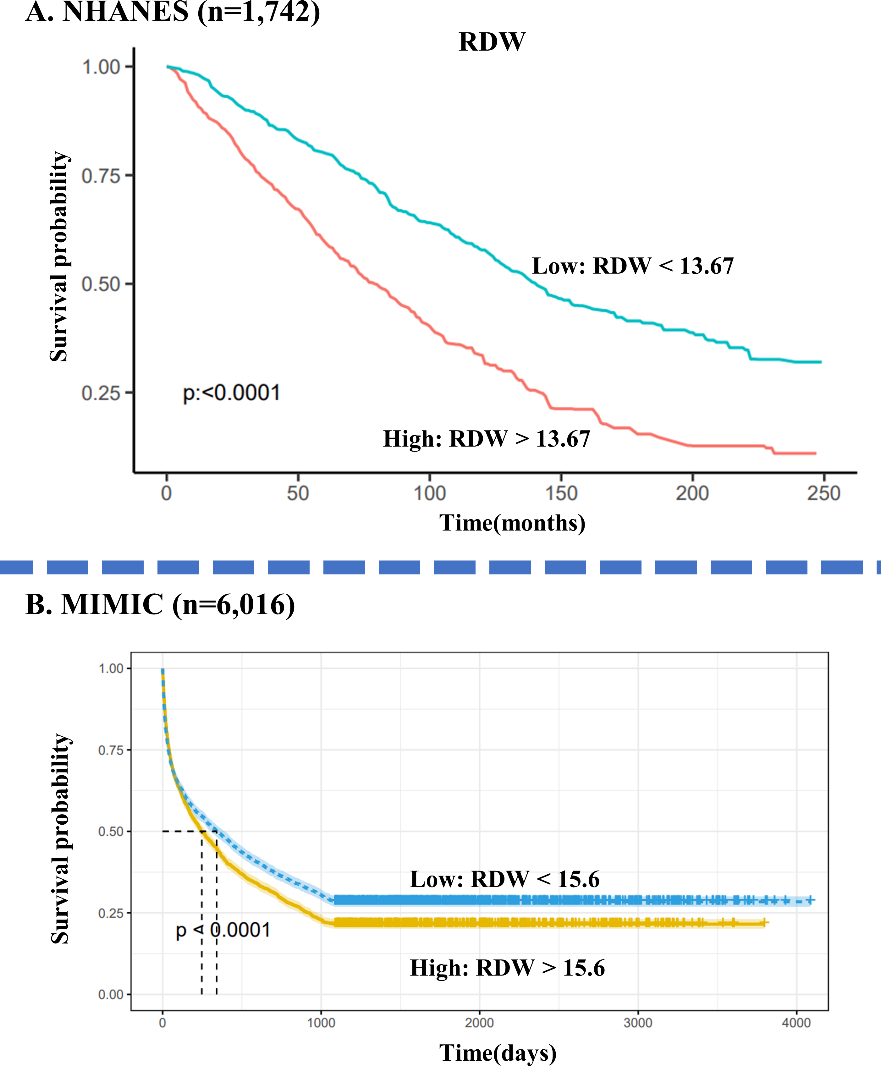

**Table S5.** The 19 quantiles mortality rate of RAR, ALB, and RDW.

| Quantile | RAR | | | | ALB | | | | RDW | | | |
| --- | --- | --- | --- | --- | --- | --- | --- | --- | --- | --- | --- | --- |
|  | value | All CHF | Dead CHF | Mortality rate | value | All CHF | Dead CHF | Mortality rate | value | All CHF | Dead CHF | Mortality rate |
| Q1 | 3.35 | 314 | 194 | 0.62 | 2 | 368 | 321 | 0.87 | 12.8 | 341 | 230 | 0.67 |
| Q2 | 3.67 | 322 | 220 | 0.68 | 2.3 | 286 | 243 | 0.85 | 13.4 | 299 | 213 | 0.71 |
| Q3 | 3.88 | 317 | 206 | 0.65 | 2.5 | 389 | 330 | 0.85 | 13.8 | 403 | 273 | 0.68 |
| Q4 | 4.03 | 309 | 213 | 0.69 | 2.6 | 284 | 219 | 0.77 | 14.1 | 252 | 183 | 0.73 |
| Q5 | 4.2 | 330 | 231 | 0.7 | 2.7 | 275 | 206 | 0.75 | 14.3 | 381 | 269 | 0.71 |
| Q6 | 4.36 | 314 | 223 | 0.71 | 2.8 | 306 | 227 | 0.74 | 14.6 | 361 | 264 | 0.73 |
| Q7 | 4.52 | 305 | 221 | 0.72 | 2.9 | 316 | 232 | 0.73 | 14.85 | 234 | 183 | 0.78 |
| Q8 | 4.69 | 329 | 248 | 0.75 | 3 | 359 | 282 | 0.79 | 15.1 | 363 | 259 | 0.71 |
| Q9 | 4.86 | 310 | 222 | 0.72 | 3.1 | 398 | 317 | 0.8 | 15.4 | 320 | 231 | 0.72 |
| Q10 | 5.03 | 314 | 229 | 0.73 | 3.2 | 372 | 288 | 0.77 | 15.7 | 312 | 244 | 0.78 |
| Q11 | 5.24 | 324 | 240 | 0.74 | 3.3 | 345 | 243 | 0.7 | 16 | 296 | 236 | 0.8 |
| Q12 | 5.43 | 306 | 224 | 0.73 | 3.4 | 346 | 255 | 0.74 | 16.3 | 278 | 207 | 0.74 |
| Q13 | 5.65 | 322 | 251 | 0.78 | 3.5 | 348 | 256 | 0.74 | 16.7 | 340 | 267 | 0.79 |
| Q14 | 5.87 | 318 | 261 | 0.82 | 3.6 | 336 | 230 | 0.68 | 17 | 291 | 216 | 0.74 |
| Q15 | 6.16 | 307 | 243 | 0.79 | 3.7 | 278 | 200 | 0.72 | 17.5 | 321 | 252 | 0.79 |
| Q16 | 6.5 | 325 | 274 | 0.84 | 3.8 | 250 | 167 | 0.67 | 18 | 305 | 253 | 0.83 |
| Q17 | 7.02 | 318 | 264 | 0.83 | 3.9 | 211 | 140 | 0.66 | 18.7 | 301 | 238 | 0.79 |
| Q18 | 7.68 | 315 | 268 | 0.85 | 4 | 287 | 192 | 0.67 | 19.7 | 305 | 234 | 0.77 |
| Q19 | 9.1 | 317 | 277 | 0.87 | 4.3 | 262 | 161 | 0.61 | 21.7 | 313 | 257 | 0.82 |
